# Impact of telehealth nutrition therapy on costs and utilization in type 2 diabetes & obesity

**DOI:** 10.1101/2025.11.09.25339829

**Authors:** Priya V. Shanmugam, Rebecca N. Adams, Shaminie J. Athinarayanan, Adam J. Wolfberg, Jeromie Ballreich

## Abstract

**Importance:** Type 2 diabetes and obesity drive substantial morbidity and spending. Rigorous evidence on the impacts of digitally delivered lifestyle interventions on healthcare cost and utilization are critical to assessing their value.

**Objective:** Determine the impact of a telehealth-delivered individualized nutrition therapy (INT) program on per-member-per-month (PMPM) total cost of care and utilization over one and two years.

**Design:** Retrospective propensity score matched difference-in-differences analysis of cost and utilization outcomes over the study period of January 2016–March 2025.

**Setting:** US adults participating in a telehealth T2D and obesity management program.

**Participants:** Enrolled in INT for >1 day or had PCP visit for T2D or obesity during study period; had ≥12 months pre-index and ≥90 days post-index claims coverage (allowing ≤30-day gaps). Final matched sample: 6,580 participants and 6,580 controls (per arm: 3,819 with type 2 diabetes and 2,761 with obesity).

**Exposures:** Telehealth-delivered, continuous care integrating individualized carbohydrate-reduced nutrition support, clinician-guided medication management, health coaching, and remote biometric monitoring.

**Main Outcomes and Measures:** Outcomes included PMPM allowed inpatient, outpatient, and prescription medication costs, inpatient, emergency department, primary care, cardiology, and endocrinology visits. For the T2D cohort, PMPM spending and proportion of days covered for each T2D medication.

**Results:** Among 3,819 adults with T2D and 2,761 adults with obesity, program participation was associated with $240 and $256 PMPM reductions in the total cost of care at 12 months (−$230 and $189 over 24 months, all P<.001). In the T2D cohort, savings were driven by deprescription of SGLT2 inhibitors (66.8% reduction in PMPM cost), sulfonylureas (51.7%), insulin (43.9%), and GLP-1s (32.2%). In the obesity cohort, reductions accrued across inpatient, outpatient and prescription medication settings.

**Conclusions and Relevance:** In this large, real-world analysis, a nutrition-first digital care model was associated with sustained reductions in cost and utilization over 12–24 months, with immediate prescription medication cost reductions in the cohort with T2D and broader savings in the cohort with obesity. Together with prior clinical evidence, these findings suggest alignment of clinical effectiveness and cost reductions of a telehealth-delivered lifestyle intervention.

**Key points:** *Question:* Is participation in a telehealth-delivered individualized nutrition therapy program associated with changes in healthcare costs and utilization among adults with type 2 diabetes (T2D) or obesity?

*Findings:* In this retrospective cohort study of 13,000 matched adults, participation was associated with lower total cost of care at one year (−$240 and −$256 for adults with T2D and obesity respectively) and over two years (T2D: -$230; obesity: $-189). Among adults with T2D, reductions were driven by inpatient costs and all T2D medications, including GLP-1s. Among adults with obesity, reductions occurred across inpatient, outpatient, and prescription medication costs.

*Meaning:* Telehealth-delivered nutrition therapy may reduce healthcare spending for adults with T2D or obesity.

## Introduction

Diabetes and obesity represent major interrelated public health challenges in the United States, impacting over 40 million^10^ and 100 million^9^ Americans respectively. Diabetes and obesity directly lead to substantial morbidity and mortality and increase the risk of myriad associated conditions including cancer, kidney disease, and cardiovascular disease.^18,41^ These syndemic conditions further create a staggering economic burden, with combined direct healthcare costs exceeding $500 billion annually.^36,47^

Despite significant advances in pharmacologic treatment options for both diabetes and obesity, including GLP-1 receptor agonists, real-world effectiveness is constrained by adverse effects, polypharmacy risks, coverage and affordability barriers that limit initiation and persistence.^2,5,20,21,23,27,30,35,39,40^ Moreover, even under guideline-concordant therapy, substantial residual cardiometabolic risk remains.^19^ Against a backdrop of rising pharmacy spend and payers’ sharpened focus on net budget impact, there is an urgent need to identify effective alternatives with tangible cost impacts. Intensive lifestyle intervention programs are effective^50^ ^28^ but limited by real-world clinical settings not designed for sustained behavior change, including limited access to registered dieticians and infrequent touchpoints.^1,14–16^ Digital health platforms now serve millions of patients, representing a paradigm shift in chronic condition management^34,11^ Wrap-around care models address gaps by delivering continuous, between-visit support integrating clinician-guided medication management, behavioral coaching, and timely specialist input. One such model is Virta Health’s Individualized Nutrition Therapy (INT) program, which is a telehealth-delivered, continuous remote care model integrating individualized carbohydrate-reduced nutrition support, clinician-guided medication management, and remote biomarker monitoring. In a prospective, controlled trial of the INT, adults with type 2 diabetes (T2D) experienced a 1.3% reduction in HbA1c and 12% weight loss at 1 year, while 94% of insulin users reduced or stopped insulin and sulfonylureas were completely eliminated.^25^ Two-year outcomes showed a durable reduction in HbA1c (0.9%) and weight (10%) with medication deintensification, translating into higher remission compared with usual care.^3^

Evidence on the total cost and utilization impact of digital wraparound care models – and thus, their comprehensive value proposition as a complement to traditional chronic condition management – is limited. This study evaluates the INT’s impacts on total cost of care (TCOC) and healthcare utilization using a retrospective claims-based analysis of over 13,000 patients over one and two years, leveraging a propensity score matched differences-in-differences (DID) study design to deliver rigorous evidence of the program’s causal impacts.

## Methods

### Study design

We conducted a retrospective matched cohort study using a difference-in-differences design to estimate the impact of digitally delivered wraparound nutritional care offered as a complement to traditional primary care-based chronic condition management (UC) compared to UC alone.

### Data sources

The study combined administrative data from the INT program with longitudinal claims data from the Komodo Healthcare Map, a nationally representative database of open and closed medical and prescription claims for over 300 million unique patients across commercial, Medicare and Medicaid plans.^31^ Datavant privacy-preserving record linkage allows for identification of unique records across payers. The study used two extracts from the Komodo Healthcare Map for the study period of January 1, 2016 through August 20, 2025. The first contained claims for INT participants, and the second contained claims for 2.4 million patients that did not participate in INT and had either two T2D or two obesity diagnosis codes >30 days apart during the study period. The study investigators did not have access to the source data used to create the extracts. Only closed claims were used for the analysis, as described in Appendix 1. Clinic data includes program fees, participation length, baseline HbA1c and weight.

All records were deidentified and compliant with United States patient confidentiality requirements, including the Health Insurance Portability and Accountability Act of 1996. This study followed the Strengthening the Reporting of Observational Studies in Epidemiology (STROBE) and RECORD-PE guidelines for observational cohort studies.^17,32^

### Study population

We compared two populations: (1) patients who enrolled in the INT between January 2017 and March 2025 (the INT arm) (2) patients who had a primary care (PCP) visit with T2D or obesity as a primary diagnosis but did not enroll in INT during the study period (the UC arm). INT and UC patients’ index date was defined as the first day of the month of INT registration or the PCP visit. The baseline period was defined as the year preceding the index date.

Patients with obesity but without T2D are treated in the INT’s Sustainable Weight Loss (SWL) track, while patients with T2D, regardless of obesity, participate in the Diabetes Reversal (DR) track. We refer to INT patients in the DR track, and UC patients with a PCP visit for T2D management, as the “cohort with T2D”. The cohort with obesity includes INT patients in the SWL track and UC patients with a PCP visit for obesity management, reflecting their primary clinical indications.

Patients with type 1 diabetes, heart failure, stage 4+ chronic kidney disease, end-stage renal disease, or acute psychosis during the baseline year were ineligible for the INT and excluded from both UC and INT arms. Patients without continuous closed claims during the baseline year or 90 days after index, allowing for gaps of up to 30 days, those aged <18 years at registration, or those with cancer or pregnancy during the baseline year were further excluded. In the T2D cohort, we excluded individuals without a baseline T2D diagnosis. In the cohort with obesity, we excluded individuals without a baseline obesity diagnosis, or with a diagnosis of T2D or use of T2D-indicated medications beyond GLP-1s and metformin. Figure 1 summarizes the study sample construction.

**Figure 1.**
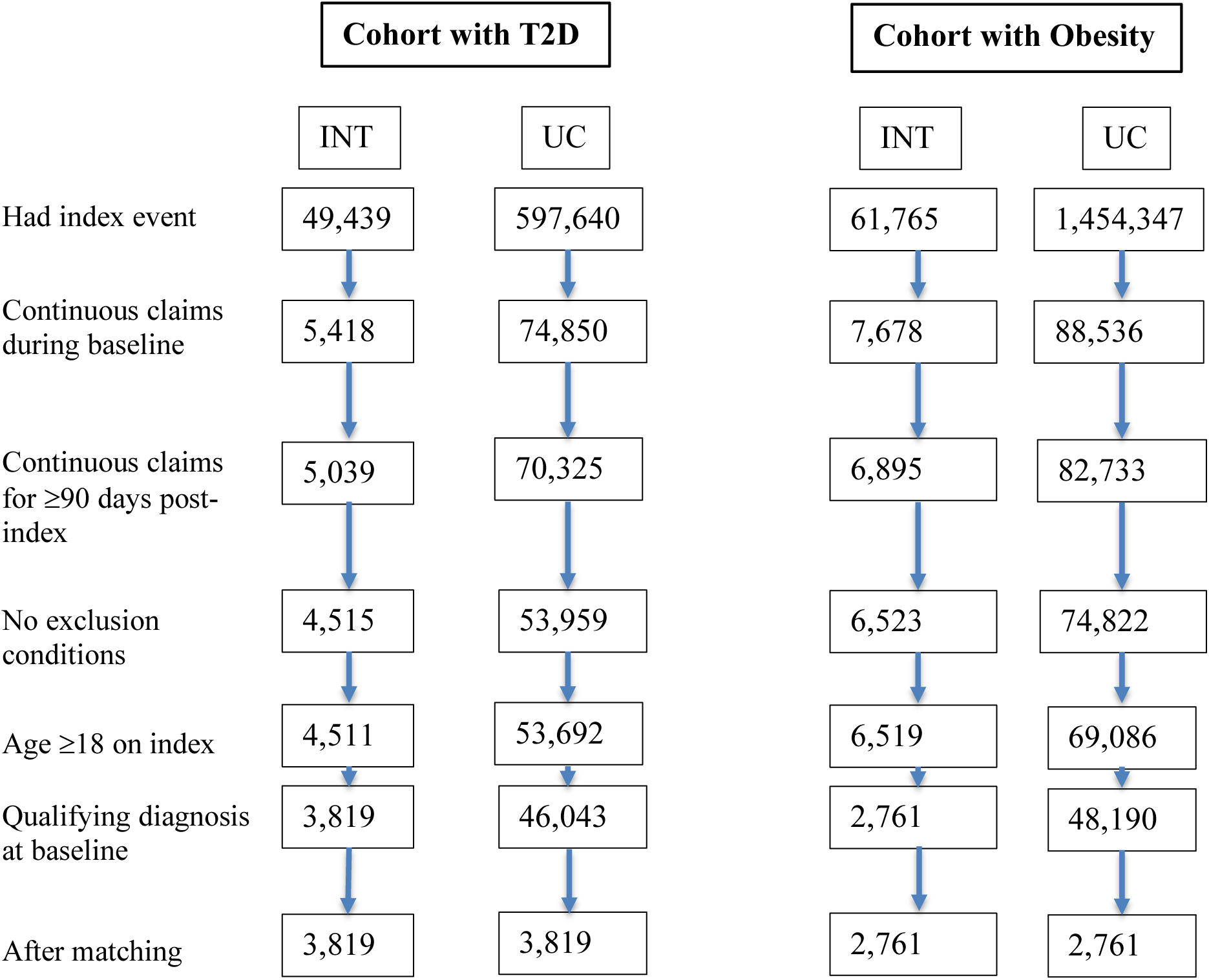
Study sample construction flowchart. **Note:** The continuous claims coverage period was defined as the period between the first and last day of closed-source medical and pharmacy claims data availability, allowing gaps ≤30 days, as detailed in Appendix 1. The index event was the first day of the month of either INT registration (for the INT arm) or a PCP visit with either T2D or obesity as a primary diagnosis (for the UC arm). The baseline period was defined as the 365 days preceding the index date. Exclusion conditions were defined by the presence of an ICD-10-CM code indicating Type 1 Diabetes, CKD Stages 4+ or ESRD, heart failure, acute psychosis, pregnancy or cancer during the baseline period. The cohort with T2D was required to have a diagnosis of T2D during baseline. The cohort with obesity was required to have a diagnosis of obesity and no diagnosis of T2D or use of SGLT2-i, sulfonylureas, thiazolidinediones, or DPP4s during baseline. **Abbreviations:** T2D: Type 2 diabetes; INT: Individualized Nutrition Therapy; UC: usual care; PCP: primary care physician; ICD-10-CM: International Classification of Diseases Volume 10 Condition Manual; CKD: chronic kidney disease; ESRD: end-stage renal disease; SGLT2-i: Sodium-glucose cotransporter-2 inhibitors; DPP-4s: dipeptidyl peitidase-4.

### Exposure

The key exposure was enrollment in the INT. Comparator patients received UC and did not participate in INT during the study period. This active-comparator design isolates the impact of INT participation by comparing participants with patients initiating primary care, reducing confounding due to motivation or engagement.

Patients are eligible for the INT if their health plan offers the program and if they have T2D, prediabetes, and/or obesity. Patients receive limited advertising about the INT and participate at no cost. Participation costs are paid by the health plan on an enrolled member-month basis.

INT participants receive telehealth-delivered continuous remote care from licensed physicians and nurse practitioners integrating individualized very low-carbohydrate nutrition support (targeting generally <30g/day), medication management, health coaching, peer support, and regular biometric feedback via a mobile app. Medication management consists of guideline-informed initiation, titration, and deprescription of diabetes- and weight-related medications based on monitoring of glycemic control, weight, and other clinical indicators, consistent with standard of care. Appendix 2 reports participation measures for the INT arm.

### Outcomes

We identified ten outcomes relevant to the management of T2D and obesity. Cost outcomes were measured per member per month (PMPM) and included costs of inpatient care, outpatient care, prescription medications, and total costs, defined as the sum of the previous three components.

Costs excluded INT program fees, which are not captured in claims. INT costs were calculated using administrative data on monthly program prices scaled by enrollment length. Utilization outcomes were reported per 1,000 members per month (PKMPM) and included inpatient visits, emergency department (ED) visits, and outpatient evaluation and management visits with primary care providers (PCPs), endocrinologists, and cardiologists.

For the T2D cohort, nine additional cost outcomes included costs for each T2D medication (metformin, thiazolidinediones, GLP-1s, SGLT2is, sulfonylureas, DPP4s, and insulin) separately, all T2D medications combined, and all non-T2D medications combined. Seven additional utilization outcomes included the proportion of days covered for each T2D medication. All outcomes were measured through the end of each patient’s closed claims coverage period, up to twenty-four months post-index.

### Covariates

The index date was the first day of the month of registration for INT (INT arm) or PCP T2D or obesity management visit (UC arm). Baseline covariates included age at index date, sex, race, comorbidities (high cholesterol, hypertension, cardiovascular disease, chronic kidney disease, liver disease, and smoking), and baseline medication use (all T2D-indicated medications and lipid- and blood pressure-lowering medications). Comorbidities and prescription medication use during the baseline year were identified by the presence of at least one claim with a corresponding ICD-10 code (Appendix 3) or medication name (Appendix 4).

### Statistical analysis

#### Matched control group construction

For the cohorts with T2D and obesity separately, matched control groups were constructed using 1:1 nearest neighbor matching on a propensity score estimated based on sex, race, five-year age groups, payer type, US region, binary indicators for baseline comorbidities and prescription medication use, and categorical indicators of baseline inpatient, outpatient, and prescription medication cost quartiles. For the cohort with obesity, the propensity score included binary indicators of baseline use of GLP-1s, other antiobesity medications, and metformin. For the cohort with T2D, the propensity score included baseline obesity and use of each T2D-indicated medication. Matching was conducted using the MatchIt package in R.^26^

#### Estimation framework

We estimated difference-in-differences regression models including indicators for INT participation, the post-index period, and their interaction. Individual and calendar-month fixed effects were included to account for time-invariant individual characteristics and secular trends. The interaction term estimated the differential change in outcomes associated with INT participation. Standard errors were clustered at the individual level.

The estimation sample was limited to the twelve months prior to, and twelve or twenty-four months following, each patient’s index date. We estimated the model for the cohort with T2D and cohort with obesity separately. For the cohort with T2D, we also estimated the model for the secondary outcomes specified above.

In this study, individuals in the comparison arm initiate a comparator intervention (PCP-based chronic condition management). In both the intervention (INT) and usual care (UC) arms, the post-period is defined relative to each patient’s initiation date. Accordingly, we used a classical 2×2 difference-in-differences (DID) specification with indicators for study arm, post-period, and their interaction. We did not use a traditional two-way fixed-effects (TWFE) DID model, in which the comparison group never receives a comparator treatment and therefore lacks an analogous index date and pre-post timing.^22^

The DID model’s key assumption is that in the absence of treatment, outcomes would have evolved similarly over time in the INT and UC arms. We assessed parallel trends for each outcome using (1) tests of differential baseline linear trends and (2) visual assessment of pre-intervention trends using an event-study specification estimating monthly coefficients relative to the month before index.

## Results

### Baseline sample characteristics

6,580 INT and 94,233 UC patients met the study eligibility criteria. The matched study sample contained 3,819 patients per arm (cohort with T2D) and 2,761 patients per arm (cohort with obesity). Table 1 shows the baseline characteristics of each cohort. The cohort with T2D averaged 55 years old at index, 52% male, and 49% white. The cohort with obesity averaged 48 years old, 29% male and 52% white. Baseline medications and comorbidities were comparable between the INT and UC arms. Appendices 5 and 6 describe the characteristics of the INT and UC arms and propensity score distribution before and after matching for each cohort.

**Table 1.**
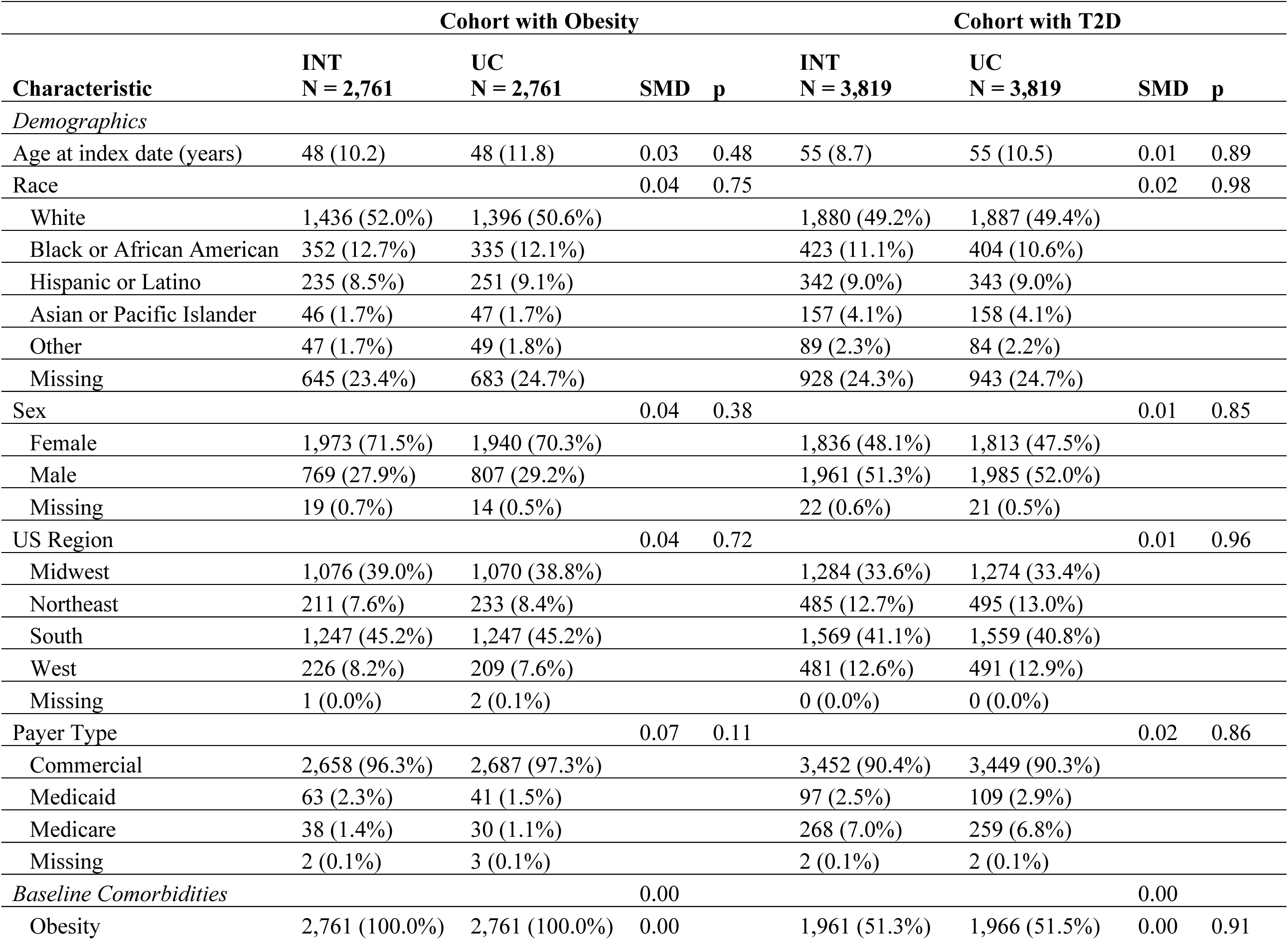
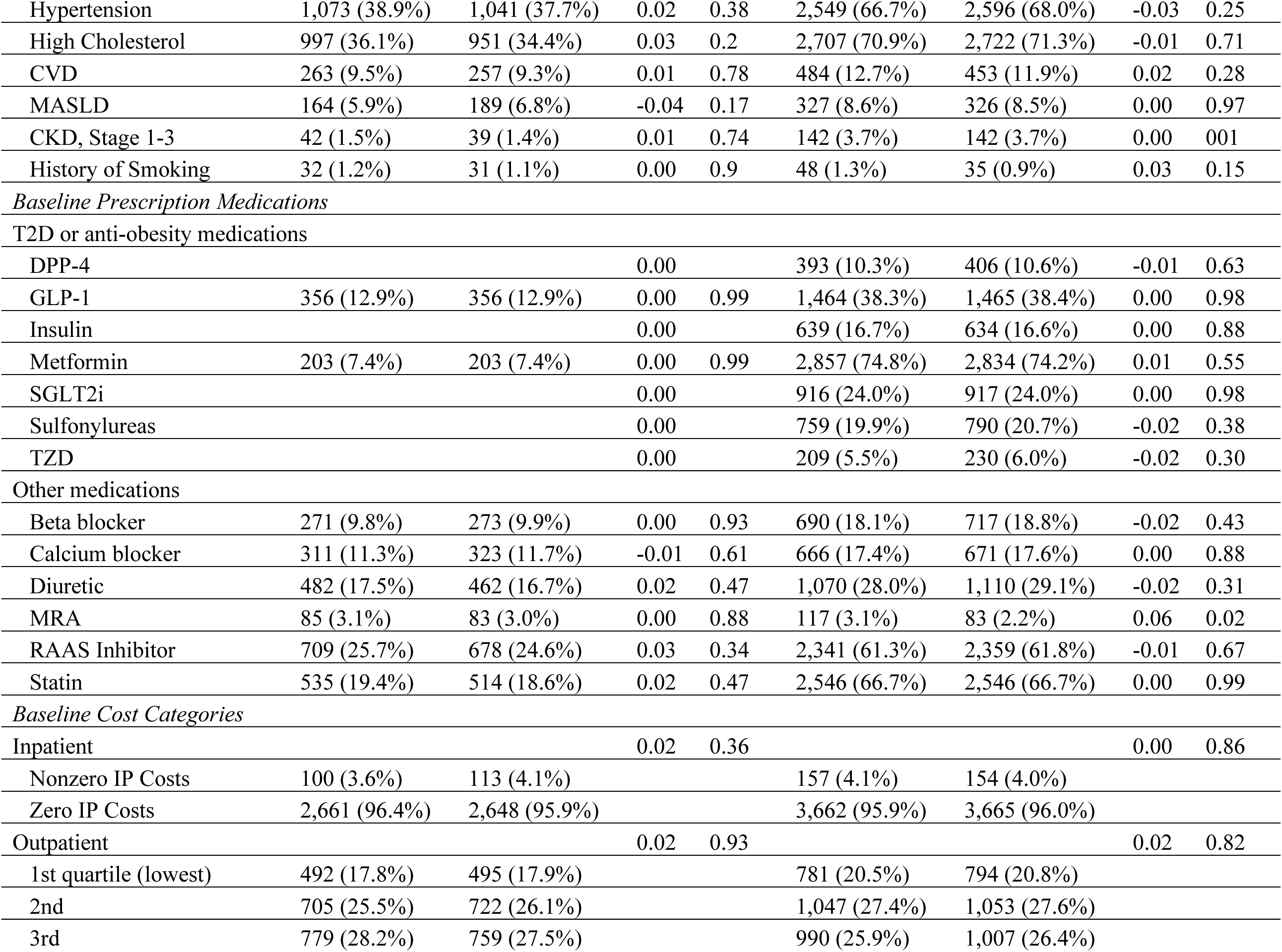
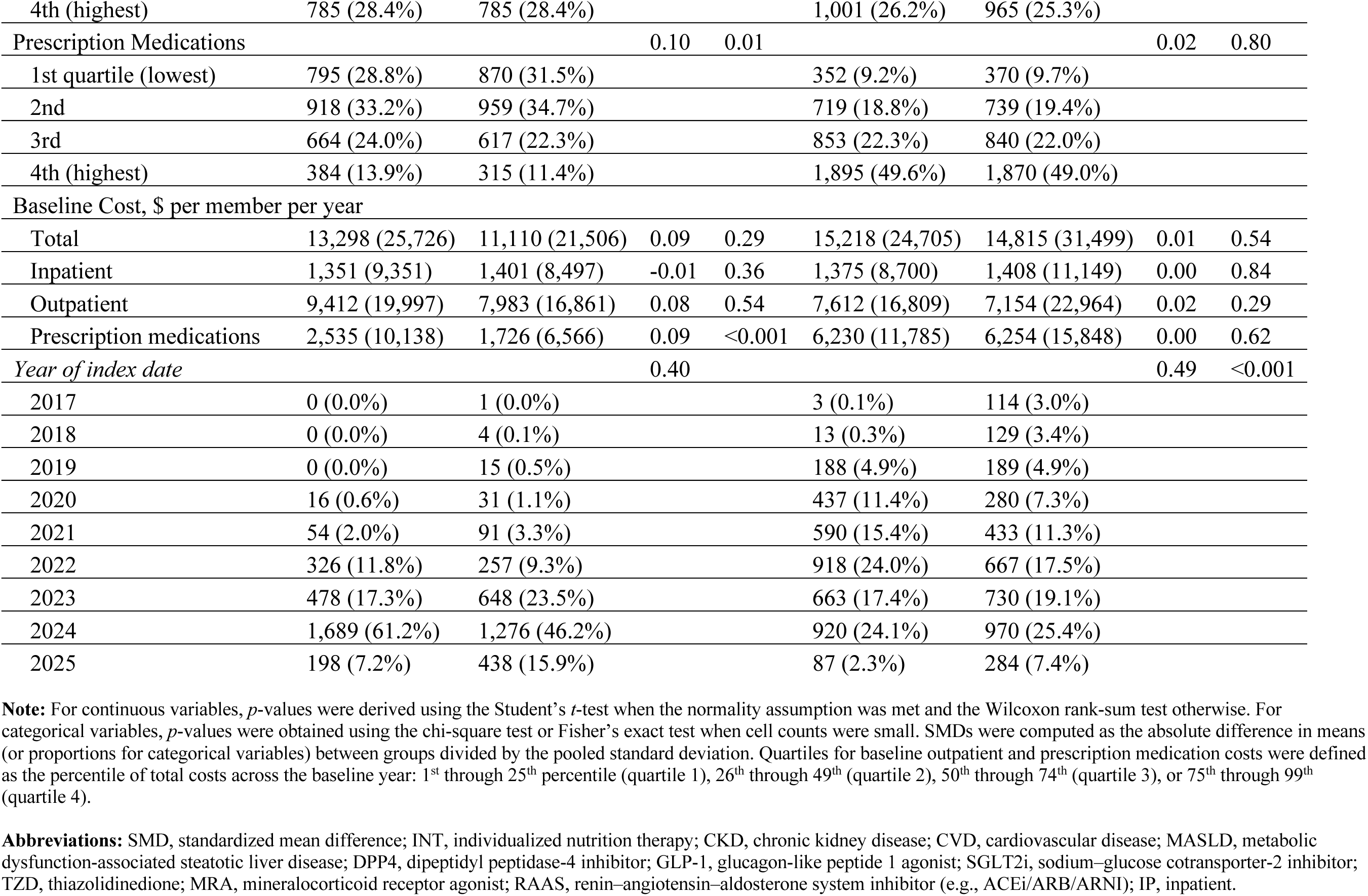
Study sample characteristics for matched cohorts with obesity and T2D.

### Impacts

Table 2 reports DID estimates by cohort. The intervention was associated with significantly lower healthcare utilization and costs in each cohort. Figure 2 reports event-study DID estimates for the twelve pre- and post-intervention months, confirming the baseline parallel trends assumption.

**Figure 2.**
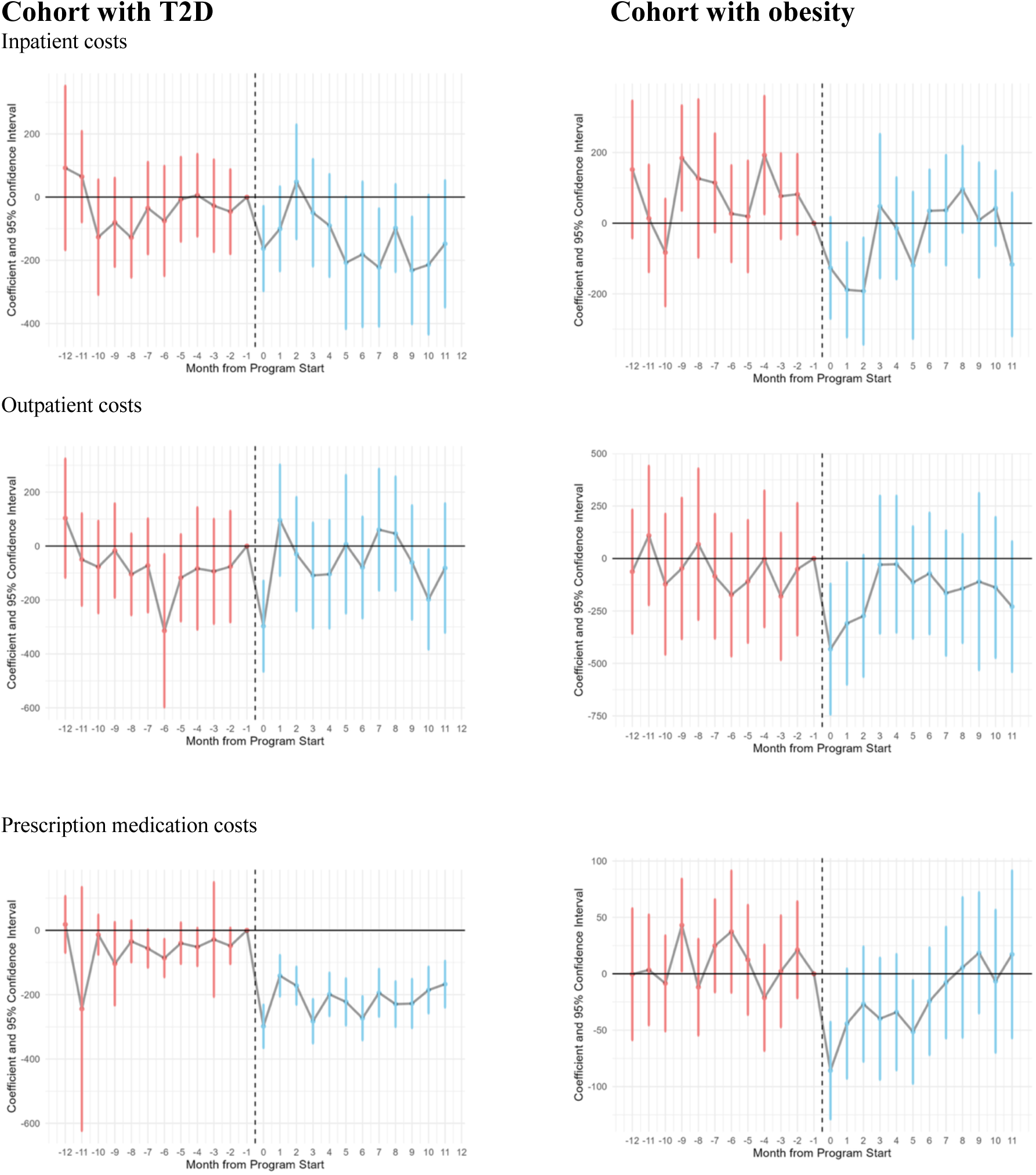
Event-study DID estimates for key outcomes. **Note:** The figure plots coefficients for the interaction terms between INT participation and each month indicator for the twelve months before and after the index date, with the month prior to the index date as the reference category. Month 0 is the first month that patients either register for the INT or have a PCP visit for T2D/obesity management. Blue estimates indicate treated months; red estimates indicate pre-treatment months. Estimates are derived from a regression model of individual monthly outcomes adjusted for individual and calendar-month fixed-effects. Standard errors are clustered at the member level. **Abbreviations:** DID, differences-in-differences; T2D, type 2 diabetes; INT, individualized nutrition therapy; PCP, primary care physician.

**Table 2.** DID estimates of cost and utilization impacts for cohorts with T2D and obesity.

| Cohort with T2D | One year |  | Two years |  | Baseline mean |  | *p*, baseline trend difference |
| --- | --- | --- | --- | --- | --- | --- | --- |
|  | DID Estimate | 95% CI | DID Estimate | 95% CI | UC | INT |  |
| *Costs, \$ per member per month* | | | | | | | |
| Total | -240.09*** | (-357.17, -123.01) | -230.25*** | (-340.36, -120.14) | 1076.62 | 1139.61 | 0.74 |
| Inpatient | -97.97** | (-161.74, -34.2) | -92.77** | (-150.16, -35.38) | 115.54 | 111.45 | 0.87 |
| Outpatient | 18.28 | (-53.23, 89.79) | 4.91 | (-61.34, 71.16) | 418.57 | 489.11 | 0.69 |
| All Prescription Medications | -160.39*** | (-210.43, -110.35) | -142.40*** | (-194.13, -90.67) | 542.51 | 539.06 | 0.28 |
| T2D-related | -145.16*** | (-161.7, -128.62) | -135.71*** | (-152.15, -119.27) | 352.02 | 359.37 | 0.26 |
| Non-T2D-related | -15.23 | (-62.04, 31.58) | -6.69 | (-55.33, 41.95) | 190.49 | 179.69 | 0.19 |
| Joint $\chi^2$ test of baseline trend difference | | | | | | | 0.53 |
| *Utilization, visits per 1000 members per month* |  |  |  |  |  |  |  |
| Inpatient | -3.2*** | (-5.0, -1.4) | -3.4*** | (-5.1, -1.8) | 4.7 | 4.3 | 0.94 |
| ED | -0.02 | (-3.3, 3.3) | -0.1 | (-3.2, 3.0) | 26.0 | 19.7 | 0.47 |
| PCP | -60.6*** | (-72.4, -48.8) | -50.2*** | (-61.6, -38.7) | 228.6 | 258.9 | 0.97 |
| Cardiologist | -0.6 | (-2.7, 1.4) | -0.2 | (-2.1, 1.7) | 8.6 | 6.5 | 0.86 |
| Endocrinologist | -4.1*** | (-6.0, -2.1) | -2.9** | (-4.8, -1.0) | 11.4 | 10.1 | 0.76 |
| Joint $\chi^2$ test of baseline trend difference | | | | | | | 0.99 |
| *N member-months* | 170,202 |  | 217,789 |  |  |  |  |

| Cohort with Obesity | One year |  | Two years |  | Baseline means |  | *p*, baseline trend difference |
| --- | --- | --- | --- | --- | --- | --- | --- |
|  | DID Estimate | 95% CI | DID Estimate | 95% CI | UC | INT |  |
| *Costs, \$ per member per month (detrended)* | | | | | | | |
| Total Cost | -256.04*** | (-374.55, -137.53) | -189.35** | (-301.32, -77.38) | 780.02 | 952.64 | 0.81 |
| Inpatient | -116.59*** | (-174.83, -58.34) | -92.04** | (-146.38, -37.69) | 112.59 | 109.29 | 0.96 |
| Outpatient | -99.28* | (-191.82, -6.75) | -72.09 | (-158.35, 14.18) | 521.40 | 647.98 | 0.79 |
| All Prescription Medications | -40.17** | (-66.37, -13.97) | -25.23 | (-52.67, 2.21) | 146.02 | 195.36 | 0.98 |
| All anti-obesity | -15.48* | (-30.23, -0.73) | 4.44 | (-9.37, 18.24) | 43.38 | 7.34 | 0.84 |
| GLP-1 | -15.42* | (-30.11, -0.73) | 4.61 | (-9.149, 18.362) | 43.30 | 7.31 | 0.84 |
| non-anti-obesity | -24.69* | (-46.54, -2.85) | -29.67* | (-53.49, -5.84) | 102.64 | 188.02 | 0.94 |
| Joint $\chi^2$ test of baseline trend difference | | | | | | | 0.99 |
| *Utilization, visits per 1,000 members per month* |  |  |  |  |  |  |  |
| Inpatient | -4.8*** | (-6.8, -2.8) | -3.8*** | (-5.6, -1.9) | 4.5 | 4.5 | 0.13 |
| ED | -5.3** | (-9.2, -1.4) | -4.9* | (-8.7, -1.1) | 22.7 | 24.9 | 0.07 |
| PCP | -74.7*** | (-90.1, -59.3) | -65.6*** | (-80.1, -51.0) | 248.4 | 277.1 | 0.00** |
| Cardiologist | -2.5* | (-4.5, -0.4) | -1.9 | (-3.7, 0.02) | 7.1 | 5.0 | 0.30 |
| Endocrinologist | -2.6** | (-4.2, -1.0) | -2.1** | (-3.6, -0.6) | 4.9 | 4.0 | 0.80 |
| Joint $\chi^2$ test of baseline trend difference | | | | | | | 0.01 |
| *N member-months* | 117,029 |  | 136,471 |  |  |  |  |

**Note:** \*\*\* $p < 0.001$ , \*\* $p < 0.01$ , \* $p < 0.05$ . The table reports coefficients, standard errors, and p-values for the interaction terms between the post-intervention indicator and the indicator for INT participation. Estimates are derived from a regression model of individual monthly outcomes adjusted for individual and calendar-month fixed-effects. Standard errors are clustered at the member level. Total cost is defined as the sum of inpatient, outpatient, and prescription medication costs. The baseline trend difference is the interaction term between INT participation and a linear month variable (1 through 12) estimated using baseline year data only. A $\chi^2$ test of the joint significance of the baseline trend difference was estimated for all cost outcomes and all utilization outcomes.
**Abbreviations:** DID, differences in differences; PMPM, per member per month; T2D, type 2 diabetes; ED, emergency department; PCP, primary care physician

#### Cohort with T2D Cost

In Year 1, total health care costs were reduced by $240 PMPM (95% CI, $123–$357), corresponding to a 21.1% reduction relative to baseline. The reduction was driven by inpatient costs ($98 PMPM [95% CI, $34–$162]) and T2D-indicated medications ($145 PMPM [95% CI, $128–$161]).

Appendix 7 reports impacts by T2D medication. Cost reductions were observed across all T2D medication classes, including SGLT2 inhibitors ($54 PMPM, or a 66.8% reduction relative to baseline), GLP-1s ($59, or 32.2%), and insulin ($22; 43.9%), with smaller, statistically significant reductions for metformin, DPP-4 inhibitors, and sulfonylureas. Reductions in the proportion of days covered were also observed for all T2D medications, including SGLT2 inhibitors (−55.5%), GLP-1s (−23.3%), and insulin (−30.8%).

Follow-up averaged 10.8 months in year one and 17.0 months over two years. Over two years, total costs were reduced by $230 PMPM, indicating sustained impacts. Table 3 compares total program fees against estimated cost reductions over one and years. Per-member net savings were $929 in year one, and $1,374 over two years.

**Table 3.** Net savings estimates.

| Cohort | Year | Net price<br>(PMPM) | Months active in<br>measurement<br>period | Total<br>program cost | DID est.<br>cost<br>reduction<br>(PMPM) | Months follow-up<br>in measurement<br>period | Cost<br>reduction<br>before fees | Net savings |  |
| --- | --- | --- | --- | --- | --- | --- | --- | --- | --- |
|  |  |  |  |  |  |  |  | (PM) | ROI |
| Diabetes | 1 | 183 | 9.5 | \$1,663 | \$240 | 10.8 | \$2,592 | \$929 | 1.56 |
| | 2 | 183 | 14.6 | \$2,536 | \$230 | 17.0 | \$3,910 | \$1,374 | 1.54 |
| Obesity | 1 | 137 | 6.8 | \$897 | \$256 | 10.0 | \$2,560 | \$1,663 | 1.85 |
| | 2 | 137 | 8.6 | \$1,118 | \$189 | 13.3 | \$2,513 | \$1,395 | 1.24 |

**Note:** The net price is the per-member, per-month blended average of list prices for the Sustainable Weight Loss and Diabetes Reversal programs over the ten-year study period and are net of performance guarantee refunds and discounts. Months active indicates the number of months INT participants were registered for the program within the one- or two-year measurement period, regardless of engagement level. Months follow-up indicates the number of months INT participants had continuous closed claims data after registration within the one- or two-year measurement period. The cost reduction before fees is the DID estimate for the total cost of care scaled by the months of follow-up. Net savings are the difference between the cost reduction before fees and the total program cost.
**Abbreviations:** PMPM, per member per month; T2D, type 2 diabetes; DPP4, dipeptidyl peptidase-4 inhibitor; GLP-1, glucagon-like peptide 1 agonist; TZD, thiazolidinedione; SGLT2i, sodium-glucose cotransporter-2 inhibitor; ROI, return on investment.

#### Utilization

In Year 1, participants had 3.2 (95% CI 1.4-5.0) fewer inpatient visits, 4.1 (2.1-6.0) fewer endocrinologist visits, and 60.6 (48.8 – 72.4) fewer PCP visits per 1,000 members per month (PKMPM). Over two years, participants had reductions of 3.4 (1.8-5.1) inpatient, 2.8 (1.0-4.8) endocrinologist, and 50.2 (38.7-61.6) fewer PCP visits PKMPM, corresponding to 40.8 fewer hospitalizations per 1,000 members annually, 19.4% fewer PCP visits and 28.6% fewer endocrinologist visits relative to the INT arm’s baseline. Impacts on ED and cardiologist visits were not statistically significant.

#### Cohort with obesity Cost

In our standard DID specification, there was a significant difference in the baseline anti-obesity medication cost trend, driven entirely by a larger increase in GLP-1 costs in the UC arm. To mitigate the impact of the differential trend, for each outcome, we estimated the baseline trend difference, detrended the outcome through year two, and estimated the model on the detrended outcome. Results for unadjusted outcomes are presented in Appendix 8. Additionally, substantial attrition was observed between years one and two in the cohort with obesity: mean follow-up was 10.0 months in the first post-index year, but 13.3 months across two years.

In Year 1, total healthcare costs were reduced by $256 PMPM (95% CI, $138-375), reflecting a 29.7% reduction from baseline. The reduction was driven by inpatient ($117 PMPM [$58-175]), outpatient ($99 [$7-192]) and prescription medications costs ($40 PMPM [$14-66]).

Over two years, total costs were $189 (77-301) PMPM lower than baseline, indicating sustained cost reductions. Table 3 compares total program fees against estimated savings, suggesting $1,374 net savings in year one and $1,395 over two years.

#### Utilization

Baseline trend estimates for low-frequency utilization outcomes (e.g., inpatient or ED visits) are imprecise, and detrending these outcomes may amplify noise or lead to poorly extrapolated outcomes. We therefore present utilization outcomes unadjusted. Participants had 4.8 (95% CI 2.8-6.8) fewer inpatient, 5.3 (1.4-9.2) fewer ED, and 74.7 (59.3-90.1) fewer PCP visits PKMPM.

Additionally, participants had 2.5 (0.4-4.5) and 2.6 (0.9-4.2) fewer cardiologist and endocrinologist visits respectively. Over two years, participants had 3.8 (1.9-5.6) fewer inpatient and 65.6 (51.0-80.1) fewer PCP visits PKMPM, corresponding to 45.6 fewer hospitalizations per 1,000 members annually and 23.7% fewer PCP visits relative to baseline.

## Discussion

In this large claims-based study of 13,000 adults with T2D and obesity, participation in a clinically effective telehealth nutrition program was associated with lower total cost of care and utilization, generating approximately $1,400 per member in net savings over two years. These findings suggest that intensive digital nutrition care can achieve both clinical effectiveness and meaningful cost reductions.

Impacts of the INT differed by clinical indication in ways consistent with the care model. Among participants with T2D, savings were driven by reductions in T2D medication spending, consistent with the rapid glycemic improvements and consequent deprescription observed in prior clinical studies.^25^ Although digital lifestyle interventions have demonstrated glycemic benefits^33,12^ prior claims-based economic evaluations often report modest or short-lived savings.^13,42,46^ The observed 12- and 24-month reductions in pharmacy and inpatient spending suggest that pairing traditional chronic condition management with telehealth-delivered nutritional care results in significant cost reductions.

In the cohort with obesity, savings accrued across a broader set of cost domains, consistent with evidence that digital support can augment clinical outcomes achieved through pharmacotherapy.^37^ Economic evaluations of prevention and weight management programs remain limited and mixed, with many reporting minimal cost reductions despite clinical benefit.^4,6,8,45^ Multiple evaluations of GLP-1s, which cost $12,000–$16,000 annually per patient^38^, have shown no cost reductions within one to five years.^48,7,49,24^ The INT compared favorably, generating $1,600 per member in net savings within one year.

The study has limitations. First, attrition through year two was substantial, particularly in the obesity cohort. Conditioning on follow-up could limit the generalizability of the results, as follow-up is determined by employment duration, or generate immortal time bias if INT participation affected employment duration and insurance coverage. Moreover, while healthcare cost reduction estimates were impacted by attrition, INT participation costs were derived from non-claims sources and thus not impacted by attrition. While this asymmetry likely places downward pressure on two-year estimates, we report estimates across both timeframes, noting that they reflect both real-world insurance churn and consequent data constraints inherent to evaluating employer-sponsored programs in commercially insured populations.

Second, digitally delivered lifestyle programs are widespread: 85% of large employers reported offering disease management programs in 2023.^29^ Because such programs paid through direct employer-vendor contracts and not captured in claims data, we cannot rule out potential contamination of the UC arm, which would bias estimated effects toward zero.

Third, residual confounding is possible in a nonrandomized observational study despite propensity score matching and a DID design. We present outcome-specific and joint baseline parallel trends tests, event-study specifications, and detrended alternate specifications to mitigate the impacts of unobserved confounding.

Fourth, the study excluded 15.3% of INT participants with T2D and 57.6% with obesity who lacked qualifying claims-based diagnoses during the baseline year, consistent with known under-recording of obesity in claims data.^44^ Linked claims and EHR data are needed to assess the generalizability of these findings to the broader cohort, particularly for INT participants with BMI-defined obesity described in Appendix 9.

Fifth, the study population was largely commercially insured. While our results are consistent with evaluations of the INT within the Veterans Health Administration,^43^ broader evidence is warranted given recent expansions in Medicare reimbursement for digitally delivered care programs.

## Conclusion

In this claims-based, real-world evaluation leveraging causal inference methods, enrollment in a telehealth nutritional care program was associated with significant reductions in healthcare utilization and costs over one and two years among adults with T2D and obesity. Reductions were driven by decreases in inpatient and pharmacy spending in the cohort with T2D and decreases in all spending categories in the cohort with obesity. These results suggest that telehealth-delivered nutritional care may be an effective method to improve outcomes and reduce healthcare costs in populations with metabolic disease.

## Data Availability

The data underlying this article was provided by the third party, Komodo Health, under license and cannot be shared publicly. The source data for this study were licensed by Virta Health from Komodo Health and hence may not be shared publicly.

## References

1. AHA. Strategies to Address Socioeconomic and Racial and Ethnic Disparities in Chronic Diseases by Incorporating Food and Nutrition Programs into the Primary Healthcare Setting. 2022. Accessed October 22, 2025. https://www.heart.org/en/-/media/Files/About-Us/Policy-Research/Policy-Positions/Access-to-Healthy-Food/Medical-Nutrition-Therapy-Policy-Statement-2022.pdf?utm_source=chatgpt.com

2. Arnold SV, Gosch K, Kosiborod M, et al. Contemporary use of cardiovascular risk reduction strategies in type 2 diabetes. Insights from the diabetes collaborative registry. Am Hear J. 2023;263:104–111. doi:10.1016/j.ahj.2023.05.002

3. Athinarayanan SJ, Adams RN, Hallberg SJ, et al. Long-Term Effects of a Novel Continuous Remote Care Intervention Including Nutritional Ketosis for the Management of Type 2 Diabetes: A 2-Year Non-randomized Clinical Trial. Front Endocrinol. 2019;10:348. doi:10.3389/fendo.2019.00348

4. Barthold D, Chiguluri V, Gumpina R, et al. Health Care Utilization and Medical Cost Outcomes from a Digital Diabetes Prevention Program in a Medicare Advantage Population. Popul Heal Manag. 2020;23(6):414–421. doi:10.1089/pop.2019.0184

5. Barthold D, Li J, Basu A. Patient Out-of-Pocket Costs for Type 2 Diabetes Medications When Aging Into Medicare. JAMA Netw Open. 2024;7(7):e2420724. doi:10.1001/jamanetworkopen.2024.20724

6. Bilger M, Finkelstein EA, Kruger E, Tate DF, Linnan LA. The Effect of Weight Loss on Health, Productivity, and Medical Expenditures Among Overweight Employees. Méd Care. 2013;51(6):471–477. doi:10.1097/mlr.0b013e318286e437

7. Bock S, Moshfegh J, Zhang J. Weighing the Impacts of GLP-1s: Quasi-Experimental Evidence From Provider Adoption. Published online February 23, 2026. https://www.nber.org/papers/w34667

8. Brown V, Tran H, Downing KL, Hesketh KD, Moodie M. A systematic review of economic evaluations of web-based or telephone-delivered interventions for preventing overweight and obesity and/or improving obesity-related behaviors. Obes Rev. 2021;22(7):e13227. doi:10.1111/obr.13227

9. CDC. Adult Obesity Facts. 2025. Accessed September 21, 2025. https://www.cdc.gov/obesity/adult-obesity-facts/index.html

10. CDC. National Diabetes Statistics Report. 2024. Accessed September 21, 2025. https://www.cdc.gov/diabetes/php/data-research/?CDC_AAref_Val=https://www.cdc.gov/diabetes/data/statistics-report/index.html

11. CDC. National Health Statistics Report. 2024. Accessed October 22, 2025. https://www.cdc.gov/nchs/data/nhsr/nhsr205.pdf

12. Crowley MJ, Tarkington PE, Bosworth HB, et al. Effect of a Comprehensive Telehealth Intervention vs Telemonitoring and Care Coordination in Patients With Persistently Poor Type 2 Diabetes Control. JAMA Intern Med. 2022;182(9):943–952. doi:10.1001/jamainternmed.2022.2947

13. Dalal MR, Robinson SB, Sullivan SD. Real-World Evaluation of the Effects of Counseling and Education in Diabetes Management. Diabetes Spectr : A Publ Am Diabetes Assoc. 2014;27(4):235–243. doi:10.2337/diaspect.27.4.235

14. Davis AM, Vogelzang JL, Affenito SG. The Shortage of Registered Dietitians or Nutritionists with a Terminal Degree: A Call to Action for the Profession. J Acad Nutr Diet. 2023;123(4):569–575. doi:10.1016/j.jand.2023.01.003

15. Deslippe AL, Soanes A, Bouchaud CC, et al. Barriers and facilitators to diet, physical activity and lifestyle behavior intervention adherence: a qualitative systematic review of the literature. Int J Behav Nutr Phys Act. 2023;20(1):14. doi:10.1186/s12966-023-01424-2

16. Echouffo-Tcheugui JB, Chakkalakal RJ, Ali MK. Is the Current Lifestyle Modification Approach to Diabetes Prevention in the U.S. a Success? Diabetes Care. 2025;48(6):863–870. doi:10.2337/dci24-0040

17. Elm E von, Altman DG, Egger M, et al. The Strengthening the Reporting of Observational Studies in Epidemiology (STROBE) statement: guidelines for reporting observational studies. J Clin Epidemiology. 2008;61(4):344–349. doi:10.1016/j.jclinepi.2007.11.008

18. Flegal KM, Graubard BI, Williamson DF, Gail MH. Cause-Specific Excess Deaths Associated With Underweight, Overweight, and Obesity. JAMA. 2007;298(17):2028–2037. doi:10.1001/jama.298.17.2028

19. Fruchart JC, Sacks FM, Hermans MP, et al. The Residual Risk Reduction Initiative: a call to action to reduce residual vascular risk in dyslipidaemic patients. Diabetes Vasc Dis Res. 2008;5(4):319–335. doi:10.3132/dvdr.2008.046

20. Gao Y, Peterson E, Pagidipati N. Barriers to prescribing glucose-lowering therapies with cardiometabolic benefits. Am Hear J. 2020;224:47–53. doi:10.1016/j.ahj.2020.03.017

21. Gleason PP, Urick BY, Marshall LZ, Friedlander N, Qiu Y, Leslie RS. Real-world persistence and adherence to glucagon-like peptide-1 receptor agonists among obese commercially insured adults without diabetes. J Manag Care Spéc Pharm. 2024;30(8):860–867. doi:10.18553/jmcp.2024.23332

22. Goodman-Bacon A. Difference-in-differences with variation in treatment timing. J Econ. 2021;225(2):254–277. doi:10.1016/j.jeconom.2021.03.014

23. Guerci B, Chanan N, Kaur S, Jasso-Mosqueda JG, Lew E. Lack of Treatment Persistence and Treatment Nonadherence as Barriers to Glycaemic Control in Patients with Type 2 Diabetes. Diabetes Ther. 2019;10(2):437–449. doi:10.1007/s13300-019-0590-x

24. Gupta N, Babyak A, Chorbajian A, Tardio V, Ballreich J, Dasgupta K. A cost-effectiveness analysis of behavioural, pharmacological, and surgical obesity treatments in Canada. *Diabetes*, Obes Metab. 2025;27(10):5748–5760. doi:10.1111/dom.16627

25. Hallberg SJ, McKenzie AL, Williams PT, et al. Effectiveness and Safety of a Novel Care Model for the Management of Type 2 Diabetes at 1 Year: An Open-Label, Non-Randomized, Controlled Study. Diabetes Ther. 2018;9(2):583–612. doi:10.1007/s13300-018-0373-9

26. Ho DE, Imai K, King G, Stuart E. {MatchIt}: Nonparametric Preprocessing for Parametric Causal Inference},. https://cran.r-project.org/web/packages/MatchIt/citation.html

27. Johansson KS, Jimenez-Solem E, Petersen TS, Christensen MB. Increasing Medication Use and Polypharmacy in Type 2 Diabetes: The Danish Experience From 2000 to 2020. Diabetes Care. 2024;47(12):2120–2127. doi:10.2337/dc24-0011

28. Kelly JT, Law L, Guzman KRD, et al. Cost-effectiveness of telehealth-delivered nutrition interventions: a systematic review of randomized controlled trials. Nutr Rev. 2023;81(12):1599–1611. doi:10.1093/nutrit/nuad032

29. KFF. 2025 *Employer Health Benefits Survey*. 2025. Accessed February 23, 2026. https://www.kff.org/health-costs/2025-employer-health-benefits-survey/#f4063214-84af-4643-b5e9-e2fa9274aa92

30. Kirkman MS, Rowan-Martin MT, Levin R, et al. Determinants of Adherence to Diabetes Medications: Findings From a Large Pharmacy Claims Database. Diabetes Care. 2015;38(4):604–609. doi:10.2337/dc14-2098

31. KomodoHealth. The Komodo Healthcare Map. October 1, 2021. https://www.komodohealth.com/solutions/healthcare-map/

32. Langan SM, Schmidt SA, Wing K, et al. The reporting of studies conducted using observational routinely collected health data statement for pharmacoepidemiology (RECORD-PE). BMJ. 2018;363:k3532. doi:10.1136/bmj.k3532

33. Lee PA, Greenfield G, Pappas Y. The impact of telehealth remote patient monitoring on glycemic control in type 2 diabetes: a systematic review and meta-analysis of systematic reviews of randomised controlled trials. BMC Heal Serv Res. 2018;18(1):495. doi:10.1186/s12913-018-3274-8

34. Morden NE, Elswyk JV, Henk HJ, Pirrello AV, Docimo AB. UnitedHealthcare’s Approach to Digitally Enabled Health Solutions: Pragmatic Trials to Identify Real Value. NEJM Catal. 2025;6(12). doi:10.1056/cat.25.0145

35. Nau DP. Recommendations for improving adherence to type 2 diabetes mellitus therapy--focus on optimizing oral and non-insulin therapies. Am J Manag care. 2012;18(3 Suppl):S49-54.

36. Parker ED, Lin J, Mahoney T, et al. Economic Costs of Diabetes in the U.S. in 2022. Diabetes Care. 2023;47(1):26–43. doi:10.2337/dci23-0085

37. Perri MG, Shankar MN, Daniels MJ, et al. Effect of Telehealth Extended Care for Maintenance of Weight Loss in Rural US Communities. JAMA Netw Open. 2020;3(6):e206764. doi:10.1001/jamanetworkopen.2020.6764

38. Peterson-KFF. How Do Prices of Drugs for Weight Loss in the U.S. Compare to Peer Nations’ Prices? 2023. Accessed October 25, 2025. https://www.healthsystemtracker.org/brief/prices-of-drugs-for-weight-loss-in-the-us-and-peer-nations/

39. Polonsky WH, Henry RR. Poor medication adherence in type 2 diabetes: recognizing the scope of the problem and its key contributors. Patient Preference Adherence. 2016;10(0):1299–1307. doi:10.2147/ppa.s106821

40. Rodriguez PJ, Zhang V, Gratzl S, et al. Discontinuation and Reinitiation of Dual-Labeled GLP-1 Receptor Agonists Among US Adults With Overweight or Obesity. JAMA Netw Open. 2025;8(1):e2457349. doi:10.1001/jamanetworkopen.2024.57349

41. Stratton IM, Adler AI, Neil HAW, et al. Association of glycaemia with macrovascular and microvascular complications of type 2 diabetes (UKPDS 35): prospective observational study. BMJ. 2000;321(7258):405. doi:10.1136/bmj.321.7258.405

42. Strawbridge LM, Lloyd JT, Meadow A, Riley GF, Howell BL. One-Year Outcomes of Diabetes Self-Management Training Among Medicare Beneficiaries Newly Diagnosed With Diabetes. Méd Care. 2017;55(4):391–397. doi:10.1097/mlr.0000000000000653

43. Strombotne KL, Lum J, Pizer SD, Figueroa S, Frakt AB, Conlin PR. Clinical effectiveness and cost-impact after 2 years of a ketogenic diet and virtual coaching intervention for patients with diabetes. *Diabetes*, Obes Metab. 2024;26(3):1016–1022. doi:10.1111/dom.15401

44. Suissa K, Schneeweiss S, Lin KJ, Brill G, Kim SC, Patorno E. Validation of obesity-related diagnosis codes in claims data. *Diabetes*, Obes Metab. 2021;23(12):2623–2631. doi:10.1111/dom.14512

45. Sweet CC, Jasik CB, Diebold A, DuPuis A, Jendretzke B. Cost Savings and Reduced Health Care Utilization Associated with Participation in a Digital Diabetes Prevention Program in an Adult Workforce Population. J Heal Econ Outcomes Res. 2020;7(2):139–147. doi:10.36469/jheor.2020.14529

46. Turner RM, Ma Q, Lorig K, Greenberg J, DeVries AR. Evaluation of a Diabetes Self-Management Program: Claims Analysis on Comorbid Illnesses, Health Care Utilization, and Cost. J Méd Internet Res. 2018;20(6):e207. doi:10.2196/jmir.9225

47. Ward ZJ, Bleich SN, Long MW, Gortmaker SL. Association of body mass index with health care expenditures in the United States by age and sex. PLoS ONE. 2021;16(3):e0247307. doi:10.1371/journal.pone.0247307

48. Wennberg D, Coetzer H, Marr A, et al. The Real-World Costs of GLP-1 Receptor Agonist Treatment. medRxiv preprint. doi:10.1101/2025.10.24.25338255v1

49. Wing C, Cai ST, Sacks D, Simon K. DO GLP-1 MEDICATIONS PAY FOR THEMSELVES? Published online February 23, 2026. https://www.nber.org/system/files/working_papers/w34678/w34678.pdf

50. Yancy WS, Crowley MJ, Dar MS, et al. Comparison of Group Medical Visits Combined With Intensive Weight Management vs Group Medical Visits Alone for Glycemia in Patients With Type 2 Diabetes. JAMA Intern Med. 2020;180(1):70–79. doi:10.1001/jamainternmed.2019.4802

