## Supplemental Materials for "Impact of telehealth nutrition therapy on costs and utilization in type 2 diabetes & obesity"

Appendix 1. Construction of closed claims periods

The Komodo Healthcare Map contains both closed and open medical and pharmacy claims data. Dates indicating the start and end of closed medical claims and closed pharmacy claims spans are provided. For the purposes of study sample construction, we define final combined periods of closed claims data using the following steps.

We first combine overlapping medical claims spans, for which the end date of the first span falls between the start and end date of the second span. For overlapping spans, we take the earlier of the two start dates, and the later of the two end dates, as our final start and end dates. Next, we combine any medical claims spans where the end of the first span is less than thirty days from the start of the second span. We repeat the above two steps for pharmacy closed claims spans.

We then identify the set of closed pharmacy and closed medical spans that overlap, taking the later of the two start dates and the earlier of the two end dates as the start and end dates for our final closed claims period. After these steps, each patient in the sample may have multiple closed claims periods. However, because the final analytic sample requires closed claims ≥365 before and ≥90 days after each patient's index event, and each patient has a maximum of one index event, a maximum of one closed claims period will remain in the estimation sample for each person after the imposition of these inclusion criteria.

The schematic below illustrates how raw closed claims periods for medical and pharmacy claims for a single individual are combined into a final closed claims period. For simplicity, each month in this illustrative calendar is 30 days in length.

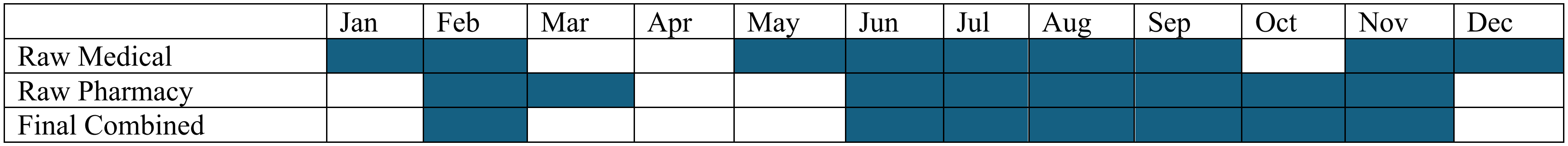

Appendix 2. Program engagement measures for cohorts with T2D and obesity

| **Measure** | **Cohort with obesity (n = 2,761)** | **Cohort with T2D (n = 3,819)** |
|---|---|---|
| *Starting HbA1c (%)* | | |
| <6.5 | 2,029 (99.6%) | 883 (28.1%) |
| 6.5-7.0 | 9 (0.4%) | 745 (23.7%) |
| 7.0-8.0 | 0 (0.0%) | 1,003 (31.9%) |
| 8.0+ | 0 (0.0%) | 515 (16.4%) |
| *Starting BMI* | | |
| <18.5 | 0 (0.0%) | 1 (0.0%) |
| 18.5-24.9 | 7 (0.4%) | 150 (5.2%) |
| 25.0-29.9 | 207 (11.0%) | 754 (26.2%) |
| 30.0-34.9 | 915 (48.5%) | 1,103 (38.3%) |
| 35.0-39.9 | 758 (40.2%) | 874 (30.3%) |
| >40.0 | 0 (0.0%) | 0 (0.0%) |
| *Ketone levels (BHB mmol/L)* | | |
| 0-0.3 | 635 (31.7%) | 1,079 (33.5%) |
| 0.3-0.5 | 711 (35.5%) | 1,082 (33.6%) |
| 0.5+ | 656 (32.8%) | 1,064 (33.0%) |
| *Time enrolled in INT program* | | |
| <6m | 914 (33.1%) | 448 (11.7%) |
| 6 to 12m | 938 (34.0%) | 933 (24.4%) |
| 12 to 24m | 666 (24.1%) | 1,221 (32.0%) |
| 24m+ | 243 (8.8%) | 1,217 (31.9%) |

**Note:** Program engagement measures are derived from administrative data from the Virta clinic and are not available for UC patients.

**Abbreviations:** T2D, type 2 diabetes; BMI, body mass index; BHB, beta-hydroxybutyrate; INT, individualized nutrition therapy.

Appendix 3. ICD-10 codesets for comorbid condition categories

| **ICD-10** | **Description** |
|---|---|
| Stroke | I63.1, I63.2, I63.3, I63.4, I63.5, I63.9 |
| Hemorrhage | I60.0, I60.1, I60.2, I60.9, I61.*, I62.* |
| Arrhythmias | I47.*, I48.*, I49.*, R00.1 |
| Ischemic Heart Disease | I20.*, I23.*, I24.*, I25.* |
| Myocardial infarction | I21.* |
| Heart failure | I50.*, I11.*, I13.* |
| Cardiac arrest | I46.2, I46.9, I97.*, R57.0 |
| Thrombosis | I80.1, I80.9, I81.*, I82.*, I26.9*, I27.82 |
| Peripheral vascular diseases | I70.*, I73.*, I74.I, I77.*, I79.* |
| Hypertension | |
| Hypertension | I10.*, I11.*, I12.*, I13.*, I15.0, I15.2, I15.9, I16.0, I16.1 |
| Diabetes (Type 2) | E11.* |
| Diabetes (Type 1) | E10.* |
| Obesity | E66.*, Z68.25, Z68.26, Z68.27, Z68.28, Z68.29. Z68.3*, Z68.4* |
| CKD Stage 1 - ESRD | N181.*, N182.*, N183.*, N184.*, N185.*, N186.* |
| Long QTc | I.4581 |
| Pregnancy | O*, Z3* |
| High Cholesterol | E78.* |
| Smoking | Z720.* |
| Cancer | C* |

Appendix 4. Prescription medication names and categories

| Drug category | Generic names |
|---|---|
| SGLT2 | Canagliflozin<br>Dapagliflozin<br>Empagliflozin<br>Ertugliflozin |
| Sulfonylureas | Glipizide<br>Glyburide<br>Glimepiride<br>Chorpropamide<br>Tolbutamide<br>Tolazamide<br>Acetohexamide |
| DPP4 | Sitagliptin<br>Saxagliptin<br>Linagliptin<br>Alogliptin<br>Vildagliptin<br>Teneligliptin |
| Thiazolidinediones | Pioglitazone<br>Rosiglitazone |
| Insulin | Insulin<br>Humulin<br>Humalog<br>Novolog |
| GLP-1 | Exenatide<br>Liraglutide<br>Lixisenatide<br>Dulaglutide<br>Semaglutide<br>Tirzepatide |
| Anticoagulant | Warfarin |

| | |
|---|---|
| | Dabigatran<br>Rivaroxaban<br>Apixaban<br>Edoxaban<br>Betrixaban |
| Antiplatelets | Anagrelide<br>Colostazol<br>Clopidogrel<br>Dipyridamole<br>Prasugrel<br>Icagrelor<br>Ticlopidine<br>Vorapaxar<br>Cangrelor |
| Statins | Atorvastatin<br>Simvastatin<br>Rosuvastatin<br>Pravastatin<br>Lovastatin<br>Fluvastatin<br>Pitavastatin<br>Ezetimibe<br>Alirocumab<br>Inclisiran |
| MRA | Eplerenone<br>Spironolactone |
| Diuretic | Bendroflumethiazide<br>Chlorothiazide<br>Chlorthalidone<br>Hydrochlorothiazide<br>Indapamide<br>Myethoclothiazide<br>Metolazone<br>Bumetanide<br>Ethacrynate sodium |

| | |
|---|---|
| | Ethacrynic acid<br>Furosemide<br>Torsemide<br>Amiloride<br>Triamterene |
| RAAS Inhibitors | Benazepril<br>Captopril<br>Enalapril<br>Fosinopril<br>Lisinopril<br>Moexipril<br>Perindopril<br>Quinapril<br>Ramipril<br>Trandolapril<br>Azilsartan<br>Candesartan<br>Eprosartan<br>Irbesartan<br>Losartan<br>Olmesartan<br>Telmisartan<br>Valsartan<br>Aliskiren |
| Beta Blockers | Atenolol<br>Betaxolol<br>Bisoprolol<br>Metoprolol tartrate<br>Metoprolol succinate<br>Nebivolol<br>Nadolol<br>Propanolol<br>Acebutolol<br>Pindolol<br>Timolol<br>Carvedilol<br>Labetalol |

| | |
|---|---|
| | Esmolol<br>Sotalol |
| Calcium Blockers | Amlodipine<br>Felodipine<br>Isradipine<br>Nicardipine<br>Nifedipine<br>Nisoldipine<br>Clevidipine<br>Nimodipine<br>Diltiazem<br>Verapamil |

Appendix 5. Characteristics of cohort with T2D before and after matching

| | Before matching | | | | After matching | | | |
|---|---|---|---|---|---|---|---|---|
| **Characteristic** | **INT N = 3,819** | **UC N = 46,043** | **SMD** | **p-val** | **INT N = 3,819** | **UC N = 3,819** | **SMD** | **p-val** |
| Demographics | | | | | | | | |
| Age at index date (years) | 55 (8.7) | 59 (12.7) | -0.37 | <0.001 | 55 (8.7) | 55 (10.5) | 0.01 | 0.89 |
| Race | | | 0.15 | <0.001 | | | 0.02 | 0.98 |
| White | 1,880 (49.2%) | 21,789 (47.3%) | | | 1,880 (49.2%) | 1,887 (49.4%) | | |
| Black or African American | 423 (11.1%) | 5,852 (12.7%) | | | 423 (11.1%) | 404 (10.6%) | | |
| Hispanic or Latino | 342 (9.0%) | 5,645 (12.3%) | | | 342 (9.0%) | 343 (9.0%) | | |
| Asian or Pacific Islander | 157 (4.1%) | 2,148 (4.7%) | | | 157 (4.1%) | 158 (4.1%) | | |
| Other | 89 (2.3%) | 1,313 (2.9%) | | | 89 (2.3%) | 84 (2.2%) | | |
| Missing | 928 (24.3%) | 9,296 (20.2%) | | | 928 (24.3%) | 943 (24.7%) | | |
| Sex | | | 0.04 | 0.05 | | | 0.01 | 0.85 |
| Female | 1,836 (48.1%) | 22,489 (48.8%) | | | 1,836 (48.1%) | 1,813 (47.5%) | | |
| Male | 1,961 (51.3%) | 23,132 (50.2%) | | | 1,961 (51.3%) | 1,985 (52.0%) | | |
| Missing | 22 (0.6%) | 422 (0.9%) | | | 22 (0.6%) | 21 (0.5%) | | |
| US Region | | | 0.41 | | | | 0.01 | 0.96 |
| Midwest | 1,284 (33.6%) | 13,370 (29.0%) | | | 1,284 (33.6%) | 1,274 (33.4%) | | |
| Northeast | 485 (12.7%) | 12,118 (26.3%) | | | 485 (12.7%) | 495 (13.0%) | | |
| South | 1,569 (41.1%) | 12,929 (28.1%) | | | 1,569 (41.1%) | 1,559 (40.8%) | | |
| Unknown | 0 (0.0%) | 5 (0.0%) | | | | | | |
| West | 481 (12.6%) | 7,621 (16.6%) | | | 481 (12.6%) | 491 (12.9%) | | |
| Payer Type | | | 0.81 | <0.001 | | | 0.02 | 0.86 |
| Commercial | 3,452 (90.4%) | 26,371 (57.3%) | | | 3,452 (90.4%) | 3,449 (90.3%) | | |
| Medicaid | 97 (2.5%) | 4,392 (9.5%) | | | 97 (2.5%) | 109 (2.9%) | | |
| Medicare | 268 (7.0%) | 14,995 (32.6%) | | | 268 (7.0%) | 259 (6.8%) | | |
| Missing | 2 (0.1%) | 285 (0.6%) | | | 2 (0.1%) | 2 (0.1%) | | |
| Baseline Comorbidities | | | | | | | | |
| Obesity | 1,961 (51.3%) | 26,965 (58.6%) | -0.15 | <0.001 | 1,961 (51.3%) | 1,966 (51.5%) | 0.00 | 0.91 |

| | | | | | | | | |
|---|---|---|---|---|---|---|---|---|
| Hypertension | 2,549 (66.7%) | 33,356 (72.4%) | -0.12 | <0.001 | 2,549 (66.7%) | 2,596 (68.0%) | -0.03 | 0.25 |
| High Cholesterol | 2,707 (70.9%) | 33,041 (71.8%) | -0.02 | 0.25 | 2,707 (70.9%) | 2,722 (71.3%) | -0.01 | 0.71 |
| MASLD | 327 (8.6%) | 4,063 (8.8%) | -0.01 | 0.58 | 327 (8.6%) | 326 (8.5%) | 0.00 | 0.97 |
| CKD, Stage 1-3 | 142 (3.7%) | 3,935 (8.5%) | -0.20 | <0.001 | 142 (3.7%) | 142 (3.7%) | 0.00 | 0.99 |
| History of Smoking | 48 (1.3%) | 1,457 (3.2%) | -0.13 | <0.001 | 48 (1.3%) | 35 (0.9%) | 0.03 | 0.15 |
| Baseline Prescription Medications | | | | | | | | |
| T2D Medications | | | | | | | | |
| DPP-4 | 393 (10.3%) | 4,225 (9.2%) | 0.04 | 0.02 | 393 (10.3%) | 406 (10.6%) | -0.01 | 0.63 |
| GLP-1 | 1,464 (38.3%) | 11,981 (26.0%) | 0.27 | <0.001 | 1,464 (38.3%) | 1,465 (38.4%) | 0.00 | 0.98 |
| Insulin | 639 (16.7%) | 7,731 (16.8%) | 0.00 | 0.93 | 639 (16.7%) | 634 (16.6%) | 0.00 | 0.88 |
| Metformin | 2,857 (74.8%) | 31,131 (67.6%) | 0.16 | <0.001 | 2,857 (74.8%) | 2,834 (74.2%) | 0.01 | 0.55 |
| SGLT2i | 916 (24.0%) | 7,240 (15.7%) | 0.21 | <0.001 | 916 (24.0%) | 917 (24.0%) | 0.00 | 0.98 |
| Sulfonylureas | 759 (19.9%) | 8,488 (18.4%) | 0.04 | 0.03 | 759 (19.9%) | 790 (20.7%) | -0.02 | 0.38 |
| TZDs | 209 (5.5%) | 2,086 (4.5%) | 0.04 | 0.01 | 209 (5.5%) | 230 (6.0%) | -0.02 | 0.30 |
| Other Medications | | | | | | | | |
| Anticoagulants | 91 (2.4%) | 2,417 (5.2%) | -0.15 | <0.001 | 91 (2.4%) | 112 (2.9%) | -0.03 | 0.14 |
| Beta blocker | 690 (18.1%) | 11,917 (25.9%) | -0.19 | <0.001 | 690 (18.1%) | 717 (18.8%) | -0.02 | 0.43 |
| Calcium blocker | 666 (17.4%) | 10,791 (23.4%) | -0.15 | <0.001 | 666 (17.4%) | 671 (17.6%) | 0.00 | 0.88 |
| Diuretic | 1,070 (28.0%) | 14,811 (32.2%) | -0.09 | <0.001 | 1,070 (28.0%) | 1,110 (29.1%) | -0.02 | 0.31 |
| MRA | 117 (3.1%) | 1,225 (2.7%) | 0.02 | 0.14 | 117 (3.1%) | 83 (2.2%) | 0.06 | 0.02 |
| RAAS Inhibitor | 2,341 (61.3%) | 27,953 (60.7%) | 0.01 | 0.47 | 2,341 (61.3%) | 2,359 (61.8%) | -0.01 | 0.67 |
| Statin | 2,546 (66.7%) | 30,089 (65.3%) | 0.03 | 0.10 | 2,546 (66.7%) | 2,546 (66.7%) | 0.00 | 001 |
| Baseline Costs | | | | | | | | |
| Inpatient | | | 0.14 | <0.001 | | | 0.00 | 0.86 |
| Nonzero IP Costs | 157 (4.1%) | 3,432 (7.5%) | | | 157 (4.1%) | 154 (4.0%) | | |
| Zero IP Costs | 3,662 (95.9%) | 42,611 (92.5%) | | | 3,662 (95.9%) | 3,665 (96.0%) | | |
| Outpatient | | | 0.03 | 0.31 | | | 0.02 | 0.82 |
| 1st (lowest) | 781 (20.5%) | 9,640 (20.9%) | | | 781 (20.5%) | 794 (20.8%) | | |
| 2nd | 1,047 (27.4%) | 11,987 (26.0%) | | | 1,047 (27.4%) | 1,053 (27.6%) | | |
| 3rd | 990 (25.9%) | 12,043 (26.2%) | | | 990 (25.9%) | 1,007 (26.4%) | | |

| | | | | | | | | |
|---|---|---|---|---|---|---|---|---|
| 4th (highest) | 1,001 (26.2%) | 12,373 (26.9%) | | | 1,001 (26.2%) | 965 (25.3%) | | |
| Prescription Medications | | | 0.18 | <0.001 | | | 0.02 | 0.8 |
| 1st (lowest) | 352 (9.2%) | 5,191 (11.3%) | | | 352 (9.2%) | 370 (9.7%) | | |
| 2nd | 719 (18.8%) | 9,135 (19.8%) | | | 719 (18.8%) | 739 (19.4%) | | |
| 3rd | 853 (22.3%) | 12,688 (27.6%) | | | 853 (22.3%) | 840 (22.0%) | | |
| 4th (highest) | 1,895 (49.6%) | 19,029 (41.3%) | | | 1,895 (49.6%) | 1,870 (49.0%) | | |
| Baseline healthcare costs, $ per member per year | | | | | | | | |
| Total | 15,218 (24,705) | 14,718 (26,151) | 0.02 | <0.001 | 15,218 (24,705) | 14,815 (31,499) | 0.01 | 0.54 |
| Inpatient | 1,375 (8,700) | 1,929 (10,597) | -0.06 | <0.001 | 1,375 (8,700) | 1,408 (11,149) | 0.00 | 0.84 |
| Outpatient | 7,612 (16,809) | 7,534 (17,751) | 0.00 | 0.43 | 7,612 (16,809) | 7,154 (22,964) | 0.02 | 0.29 |
| Prescription medications | 6,230 (11,785) | 5,255 (11,763) | 0.08 | <0.001 | 6,230 (11,785) | 6,254 (15,848) | 0.00 | 0.62 |
| Index Year | | | 0.59 | <0.001 | | | 0.49 | <0.001 |
| 2016 | 0 (0.0%) | 1,047 (2.3%) | | | 0 (0.0%) | 23 (0.6%) | | |
| 2017 | 3 (0.1%) | 2,224 (4.8%) | | | 3 (0.1%) | 114 (3.0%) | | |
| 2018 | 13 (0.3%) | 2,974 (6.5%) | | | 13 (0.3%) | 129 (3.4%) | | |
| 2019 | 188 (4.9%) | 3,340 (7.3%) | | | 188 (4.9%) | 189 (4.9%) | | |
| 2020 | 437 (11.4%) | 4,515 (9.8%) | | | 437 (11.4%) | 280 (7.3%) | | |
| 2021 | 590 (15.4%) | 6,366 (13.8%) | | | 590 (15.4%) | 433 (11.3%) | | |
| 2022 | 918 (24.0%) | 7,672 (16.7%) | | | 918 (24.0%) | 667 (17.5%) | | |
| 2023 | 663 (17.4%) | 7,803 (16.9%) | | | 663 (17.4%) | 730 (19.1%) | | |
| 2024 | 920 (24.1%) | 8,086 (17.6%) | | | 920 (24.1%) | 970 (25.4%) | | |
| 2025 | 87 (2.3%) | 2,016 (4.4%) | | | 87 (2.3%) | 284 (7.4%) | | |

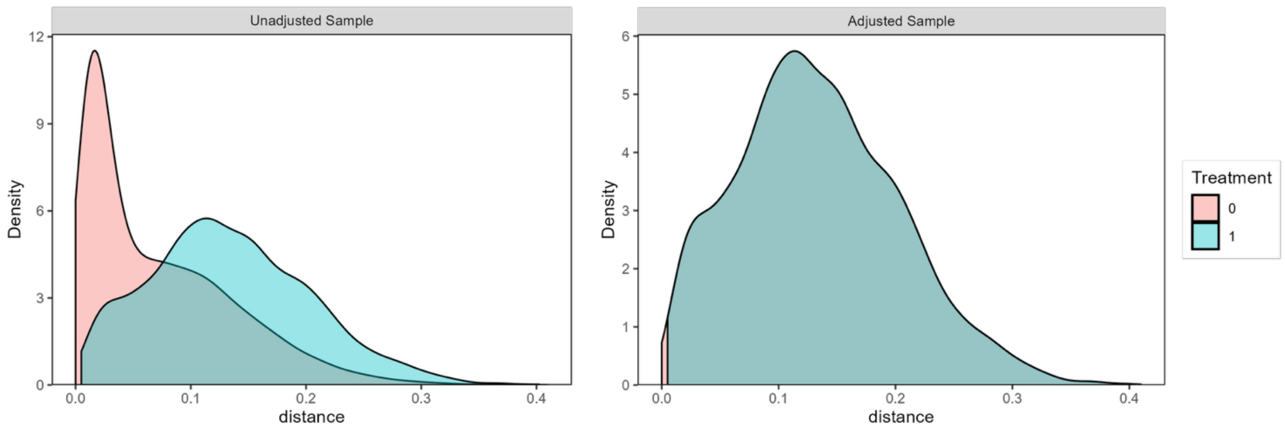

Unadjusted Sample
Adjusted Sample
Density
distance
0
3
6
9
12
0.0
0.1
0.2
0.3
0.4
1
2
4
5
Treatment
0
1

Appendix 6. Characteristics of cohort with obesity before and after matching

| | Before matching | | | | After matching | | | |
|---|---|---|---|---|---|---|---|---|
| **Characteristic** | **INT<br>N = 2,761** | **UC<br>N = 48,190** | **SMD** | **p-val** | **INT<br>N = 2,761** | **UC<br>N = 2,761** | **SMD** | **p-val** |
| Demographics | | | | | | | | |
| Age at index date (years) | 48 (10.2) | 48 (14.4) | 0.06 | <0.001 | 48 (10.2) | 48 (11.8) | 0.03 | 0.48 |
| Race | | | 0.15 | <0.001 | | | 0.04 | 0.75 |
| White | 1,436 (52.0%) | 22,647 (47.0%) | | | 1,436 (52.0%) | 1,396 (50.6%) | | |
| Black or African American | 352 (12.7%) | 5,575 (11.6%) | | | 352 (12.7%) | 335 (12.1%) | | |
| Hispanic or Latino | 235 (8.5%) | 5,200 (10.8%) | | | 235 (8.5%) | 251 (9.1%) | | |
| Asian or Pacific Islander | 46 (1.7%) | 1,116 (2.3%) | | | 46 (1.7%) | 47 (1.7%) | | |
| Other | 47 (1.7%) | 1,464 (3.0%) | | | 47 (1.7%) | 49 (1.8%) | | |
| Missing | 645 (23.4%) | 12,188 (25.3%) | | | 645 (23.4%) | 683 (24.7%) | | |
| Sex | | | 0.08 | <0.001 | | | 0.04 | 0.38 |
| Female | 1,973 (71.5%) | 32,625 (67.7%) | | | 1,973 (71.5%) | 1,940 (70.3%) | | |
| Male | 769 (27.9%) | 15,170 (31.5%) | | | 769 (27.9%) | 807 (29.2%) | | |
| Missing | 19 (0.7%) | 395 (0.8%) | | | 19 (0.7%) | 14 (0.5%) | | |
| US Region | | | 0.63 | | | | 0.04 | 0.72 |
| Midwest | 1,076 (39.0%) | 15,073 (31.3%) | | | 1,076 (39.0%) | 1,070 (38.8%) | | |
| Northeast | 211 (7.6%) | 13,062 (27.1%) | | | 211 (7.6%) | 233 (8.4%) | | |
| South | 1,247 (45.2%) | 13,121 (27.2%) | | | 1,247 (45.2%) | 1,247 (45.2%) | | |
| Unknown | 1 (0.0%) | 6 (0.0%) | | | 1 (0.0%) | 2 (0.1%) | | |
| West | 226 (8.2%) | 6,928 (14.4%) | | | 226 (8.2%) | 209 (7.6%) | | |
| Payer Type | | | 0.71 | <0.001 | | | 0.07 | 0.11 |
| Commercial | 2,658 (96.3%) | 34,691 (72.0%) | | | 2,658 (96.3%) | 2,687 (97.3%) | | |
| Medicaid | 63 (2.3%) | 6,105 (12.7%) | | | 63 (2.3%) | 41 (1.5%) | | |
| Medicare | 38 (1.4%) | 7,195 (14.9%) | | | 38 (1.4%) | 30 (1.1%) | | |
| Missing | 2 (0.1%) | 199 (0.4%) | | | 2 (0.1%) | 3 (0.1%) | | |
| Baseline Comorbidities | | | | | | | | |
| Hypertension | 1,073 (38.9%) | 19,327 (40.1%) | -0.03 | 0.19 | 1,073 (38.9%) | 1,041 (37.7%) | 0.02 | 0.38 |

| | | | | | | | | |
|---|---|---|---|---|---|---|---|---|
| High Cholesterol | 997 (36.1%) | 18,971 (39.4%) | -0.07 | <0.001 | 997 (36.1%) | 951 (34.4%) | 0.03 | 0.20 |
| CVD | 263 (9.5%) | 5,326 (11.1%) | -0.05 | 0.01 | 263 (9.5%) | 257 (9.3%) | 0.01 | 0.78 |
| MASLD | 164 (5.9%) | 3,184 (6.6%) | -0.03 | 0.17 | 164 (5.9%) | 189 (6.8%) | -0.04 | 0.17 |
| CKD, Stage 1-3 | 42 (1.5%) | 1,090 (2.3%) | -0.05 | 0.01 | 42 (1.5%) | 39 (1.4%) | 0.01 | 0.74 |
| History of Smoking | 32 (1.2%) | 1,291 (2.7%) | -0.11 | <0.001 | 32 (1.2%) | 31 (1.1%) | 0.00 | 0.9 |
| Baseline Prescription Medications | | | | | | | | |
| GLP-1 | 356 (12.9%) | 6,897 (14.3%) | -0.04 | 0.04 | 356 (12.9%) | 356 (12.9%) | 0.00 | 0.99 |
| Metformin | 203 (7.4%) | 2,874 (6.0%) | 0.06 | 0.003 | 203 (7.4%) | 203 (7.4%) | 0.00 | 0.99 |
| Anticoagulants | 58 (2.1%) | 1,362 (2.8%) | -0.05 | 0.02 | 58 (2.1%) | 50 (1.8%) | 0.02 | 0.44 |
| Beta blocker | 271 (9.8%) | 5,632 (11.7%) | -0.06 | 0.002 | 271 (9.8%) | 273 (9.9%) | 0.00 | 0.93 |
| Calcium blocker | 311 (11.3%) | 5,891 (12.2%) | -0.03 | 0.13 | 311 (11.3%) | 323 (11.7%) | -0.01 | 0.61 |
| Diuretic | 482 (17.5%) | 8,626 (17.9%) | -0.01 | 0.56 | 482 (17.5%) | 462 (16.7%) | 0.02 | 0.47 |
| MRA | 85 (3.1%) | 1,262 (2.6%) | 0.03 | 0.14 | 85 (3.1%) | 83 (3.0%) | 0.00 | 0.88 |
| RAAS Inhibitor | 709 (25.7%) | 11,641 (24.2%) | 0.04 | 0.07 | 709 (25.7%) | 678 (24.6%) | 0.03 | 0.34 |
| Statin | 535 (19.4%) | 9,302 (19.3%) | 0.00 | 0.92 | 535 (19.4%) | 514 (18.6%) | 0.02 | 0.47 |
| Baseline Cost Categories | | | | | | | | |
| Inpatient | | | 0.10 | <0.001 | | | 0.02 | 0.36 |
| Nonzero IP Costs | 100 (3.6%) | 2,781 (5.8%) | | | 100 (3.6%) | 113 (4.1%) | | |
| Zero IP Costs | 2,661 (96.4%) | 45,409 (94.2%) | | | 2,661 (96.4%) | 2,648 (95.9%) | | |
| Outpatient | | | 0.06 | 0.047 | | | 0.02 | 0.93 |
| 1st quartile (lowest) | 492 (17.8%) | 9,598 (19.9%) | | | 492 (17.8%) | 495 (17.9%) | | |
| 2nd | 705 (25.5%) | 12,301 (25.5%) | | | 705 (25.5%) | 722 (26.1%) | | |
| 3rd | 779 (28.2%) | 13,013 (27.0%) | | | 779 (28.2%) | 759 (27.5%) | | |
| 4th (highest) | 785 (28.4%) | 13,278 (27.6%) | | | 785 (28.4%) | 785 (28.4%) | | |
| Prescription Medications | | | 0.12 | <0.001 | | | 0.10 | 0.01 |
| 1st quartile (lowest) | 795 (28.8%) | 14,269 (29.6%) | | | 795 (28.8%) | 870 (31.5%) | | |
| 2nd | 918 (33.2%) | 13,679 (28.4%) | | | 918 (33.2%) | 959 (34.7%) | | |
| 3rd | 664 (24.0%) | 12,225 (25.4%) | | | 664 (24.0%) | 617 (22.3%) | | |
| 4th (highest) | 384 (13.9%) | 8,017 (16.6%) | | | 384 (13.9%) | 315 (11.4%) | | |
| Baseline costs, $ per member per year | | | | | | | | |

| | | | | | | | | |
|---|---|---|---|---|---|---|---|---|
| Total | 13,298 (25,726.0) | 11,351 (22,751.6) | 0.08 | 0.76 | 13,298 (25,726.0) | 11,110 (21,506.4) | 0.09 | 0.29 |
| Inpatient | 1,351 (9,351.3) | 1,477 (8,336.5) | -0.01 | <0.001 | 1,351 (9,351.3) | 1,401 (8,496.8) | -0.01 | 0.36 |
| Outpatient | 9,412 (19,997.5) | 7,365 (15,982.0) | 0.11 | 0.004 | 9,412 (19,997.5) | 7,983 (16,861.4) | 0.08 | 0.54 |
| Prescription medications | 2,535 (10,137.5) | 2,509 (10,207.3) | 0.00 | 0.03 | 2,535 (10,137.5) | 1,726 (6,566.0) | 0.09 | <0.001 |
| Year of index date | | | 1.2 | <0.001 | | | 0.40 | |
| 2016 | 0 (0.0%) | 1,002 (2.1%) | | | 0 (0.0%) | 0 (0.0%) | | |
| 2017 | 0 (0.0%) | 2,173 (4.5%) | | | 0 (0.0%) | 1 (0.0%) | | |
| 2018 | 0 (0.0%) | 2,878 (6.0%) | | | 0 (0.0%) | 4 (0.1%) | | |
| 2019 | 0 (0.0%) | 3,187 (6.6%) | | | 0 (0.0%) | 15 (0.5%) | | |
| 2020 | 16 (0.6%) | 3,849 (8.0%) | | | 16 (0.6%) | 31 (1.1%) | | |
| 2021 | 54 (2.0%) | 5,682 (11.8%) | | | 54 (2.0%) | 91 (3.3%) | | |
| 2022 | 326 (11.8%) | 7,284 (15.1%) | | | 326 (11.8%) | 257 (9.3%) | | |
| 2023 | 478 (17.3%) | 9,165 (19.0%) | | | 478 (17.3%) | 648 (23.5%) | | |
| 2024 | 1,689 (61.2%) | 10,129 (21.0%) | | | 1,689 (61.2%) | 1,276 (46.2%) | | |
| 2025 | 198 (7.2%) | 2,841 (5.9%) | | | 198 (7.2%) | 438 (15.9%) | | |

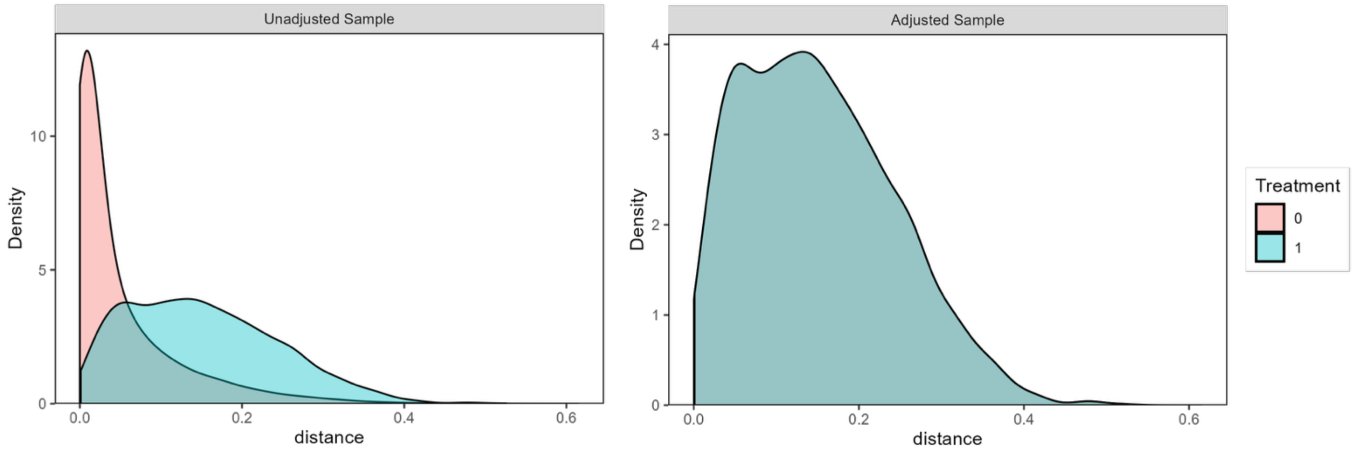

Appendix 7. DID estimates for specific T2D medications, for cohort with T2D

| T2D Medication | One year | | Two years | | Baseline Means | | Relative change |
|---|---|---|---|---|---|---|---|
| | DID Estimate | 95% CI | DID Estimate | 95% CI | UC | INT | |
| *Cost, $ per member per month* | | | | | | | |
| Metformin | -5.30*** | (-2.43, -8.17) | -5.75*** | (-2.88, -8.63) | 18.35 | 18.04 | -29.4% |
| DPP-4 | -4.77** | (-1.66, -7.88) | -4.72** | (-1.43, -8.01) | 26.20 | 26.37 | -18.1% |
| GLP-1 | -58.90*** | (-46.13, -71.67) | -50.83*** | (-38.24, -63.42) | 181.25 | 182.93 | -32.2% |
| TZD | 0.06 | (0.18, -0.06) | 0.03 | (0.15, -0.09) | 0.34 | 0.26 | 23.3% |
| Sulfonylureas | -0.38*** | (-0.3, -0.46) | -0.38*** | (-0.3, -0.46) | 0.69 | 0.73 | -51.7% |
| SGLT2i | -53.50*** | (-46.92, -60.09) | -51.32*** | (-44.64, -58) | 80.10 | 80.04 | -66.8% |
| Insulin | -22.38*** | (-17.04, -27.71) | -22.73*** | (-17.27, -28.19) | 45.09 | 51.00 | -43.9% |
| *Proportion of Days Covered, Percentage Point* | | | | | | | |
| Metformin | -0.8 | (-0.02, 0.01) | -1.7* | (-0.03, -0.002) | 56.1 | 58.6 | -1.4% |
| DPP-4 | -0.8* | (-0.02, -0.001) | -1.0* | (-0.02, -0.002) | 7.3 | 7.2 | -11.5% |
| GLP-1 | -6.3*** | (-0.08, -0.05) | -5.7*** | (-0.07, -0.04) | 25.6 | 26.8 | -23.3% |
| TZD | -0.9** | (-0.02, -0.004) | -1.1*** | (-0.02, -0.01) | 4.2 | 4.1 | -22.2% |
| Sulfonylureas | -5.6*** | (-0.07, -0.05) | -6.1*** | (-0.07, -0.05) | 15.3 | 14.8 | -37.9% |
| SGLT2i | -9.5*** | (-0.11, -0.08) | -9.7*** | (-0.11, -0.09) | 16.7 | 17.1 | -55.5% |
| Insulin | -3.9*** | (-0.05, -0.03) | -4.4*** | (-0.05, -0.04) | 11.6 | 12.6 | -30.8% |

**Note:** The table reports coefficients, standard errors, and p-values for the interaction terms between the post-intervention indicator and the indicator for INT participation. Estimates are derived from a regression model of individual monthly outcomes adjusted for individual and calendar-month fixed-effects. Standard errors are clustered at the member level. Relative changes are expressed by dividing the one-year DID estimate by the baseline mean for the INT group.

**Abbreviations:** PMPM, per member per month; T2D, type 2 diabetes; DPP4, dipeptidyl peptidase-4 inhibitor; GLP-1, glucagon-like peptide 1 agonist; TZD, thiazolidinedione; SGLT2i, sodium–glucose cotransporter-2 inhibitor.

Appendix 8. Results for unadjusted cost outcomes in cohort with obesity

| | One year | | Two years | | Baseline Means | | p-val, baseline trend |
|---|---|---|---|---|---|---|---|
| **Outcome** | **DID Estimate** | **95% CI** | **DID Estimate** | **95% CI** | **UC** | **INT** | |
| *Cost, $ per member per month* | | | | | | | |
| Total Cost | -322.94*** | (-441.44, -204.44) | -270.25*** | (-382.2, -158.3) | 780.02 | 990.92 | 0.45 |
| Inpatient | -123.99*** | (-182.24, -65.74) | -100.99*** | (-155.34, -46.64) | 112.59 | 113.53 | 0.86 |
| Outpatient | -128.90** | (-221.43, -36.37) | -107.90* | (-194.16, -21.64) | 521.40 | 664.93 | 0.59 |
| All Prescription Medications | -70.05*** | (-96.24, -43.86) | -61.36*** | (-88.79, -33.93) | 146.02 | 212.46 | 0.11 |
| All anti-obesity | -61.10*** | (-75.85, -46.35) | -50.72*** | (-64.5, -36.94) | 43.38 | 33.45 | 0.00 |
| GLP-1s | -61.03*** | (-75.72, -46.35) | -50.55*** | (-64.28, -36.82) | 43.30 | 33.41 | 0.00 |
| All non-anti-obesity | -8.95 | (-30.8, 12.9) | -10.63 | (-34.45, 13.19) | 102.64 | 179.01 | 0.31 |

**Note:** *** $p < 0.001$, ** $p < 0.01$, * $p < 0.05$. The table reports coefficients, standard errors, and p-values for the interaction terms between the post-intervention indicator and the indicator for INT participation. Estimates are derived from a regression model of individual monthly outcomes adjusted for individual and calendar-month fixed-effects. Standard errors are clustered at the member level. Total cost is defined as the sum of inpatient, outpatient, and prescription medication costs. The baseline trend difference is the interaction term between INT participation and a linear month variable (1 through 12) estimated using baseline year data only. A $\chi^2$ test of the joint significance of the baseline trend difference was estimated for all cost outcomes and all utilization outcomes.

**Abbreviations:** DID, differences in differences; PMPM, per member per month; T2D, type 2 diabetes; ED, emergency department; PCP, primary care physician

Appendix 9. Characteristics of INT participants with and without ICD-10 code

| **Characteristic** | **Without ICD-10 (N = 3,518)** | **With ICD-10 (N = 2,776)** | **SMD** | **p** |
|---|---|---|---|---|
| Demographics | | | | |
| Age at index date (years) | 48 (10.5) | 48 (10.2) | -0.06 | 0.02 |
| Race | | | 0.22 | <0.001 |
| White | 1,862 (52.9%) | 1,445 (52.1%) | | |
| Black or African American | 261 (7.4%) | 355 (12.8%) | | |
| Hispanic or Latino | 267 (7.6%) | 235 (8.5%) | | |
| Asian or Pacific Islander | 117 (3.3%) | 46 (1.7%) | | |
| Other | 63 (1.8%) | 47 (1.7%) | | |
| Missing | 948 (26.9%) | 648 (23.3%) | | |
| Sex | | | 0.17 | <0.001 |
| Female | 2,256 (64.1%) | 1,987 (71.6%) | | |
| Male | 1,244 (35.4%) | 770 (27.7%) | | |
| Missing | 18 (0.5%) | 19 (0.7%) | | |
| US Region | | | 0.27 | |
| Midwest | 1,672 (47.5%) | 1,083 (39.0%) | | |
| Northeast | 235 (6.7%) | 213 (7.7%) | | |
| South | 1,174 (33.4%) | 1,251 (45.1%) | | |
| Unknown | 0 (0.0%) | 1 (0.0%) | | |
| West | 437 (12.4%) | 228 (8.2%) | | |
| Payer Type | | | 0.05 | 0.21 |
| Commercial | 3,350 (95.2%) | 2,672 (96.3%) | | |
| Medicaid | 110 (3.1%) | 64 (2.3%) | | |
| Medicare | 55 (1.6%) | 38 (1.4%) | | |
| Missing | 3 (0.1%) | 2 (0.1%) | | |
| Baseline Comorbidities | | | | |
| Hypertension | 705 (20.0%) | 1,078 (38.8%) | -0.42 | <0.001 |
| High Cholesterol | 759 (21.6%) | 999 (36.0%) | -0.32 | <0.001 |
| CVD | 191 (5.4%) | 263 (9.5%) | -0.15 | <0.001 |

| | | | | |
|---|---|---|---|---|
| MASLD | 88 (2.5%) | 165 (5.9%) | -0.17 | <0.001 |
| CKD, Stage 1-3 | 19 (0.5%) | 42 (1.5%) | -0.10 | <0.001 |
| History of Smoking | 25 (0.7%) | 32 (1.2%) | -0.05 | 0.07 |
| Baseline Prescription Medications | | | | |
| DPP4 | 2 (0.1%) | 1 (0.0%) | 0.01 | 0.99 |
| GLP-1 | 104 (3.0%) | 358 (12.9%) | -0.37 | <0.001 |
| Insulin | 9 (0.3%) | 12 (0.4%) | -0.03 | 0.23 |
| Metformin | 126 (3.6%) | 205 (7.4%) | -0.17 | <0.001 |
| SGLT2i | 3 (0.1%) | 3 (0.1%) | -0.01 | 0.99 |
| Sulfonylureas | 1 (0.0%) | 0 (0.0%) | 0.02 | 0.99 |
| TZDs | | | 0.00 | |
| Anticoagulants | 46 (1.3%) | 59 (2.1%) | -0.06 | 0.01 |
| Beta blocker | 204 (5.8%) | 275 (9.9%) | -0.15 | <0.001 |
| Calcium blocker | 275 (7.8%) | 314 (11.3%) | -0.12 | <0.001 |
| Diuretic | 359 (10.2%) | 488 (17.6%) | -0.21 | <0.001 |
| MRA | 82 (2.3%) | 85 (3.1%) | -0.05 | 0.07 |
| RAAS Inhibitor | 595 (16.9%) | 713 (25.7%) | -0.22 | <0.001 |
| Statin | 495 (14.1%) | 537 (19.3%) | -0.14 | <0.001 |
| Baseline Cost Categories | | | | |
| Inpatient | | | 0.13 | <0.001 |
| Nonzero IP Costs | 58 (1.6%) | 105 (3.8%) | | |
| Zero IP Costs | 3,460 (98.4%) | 2,671 (96.2%) | | |
| Outpatient | | | 0.54 | <0.001 |
| 1st quartile (lowest) | 1,413 (40.2%) | 494 (17.8%) | | |
| 2nd | 857 (24.4%) | 706 (25.4%) | | |
| 3rd | 681 (19.4%) | 783 (28.2%) | | |
| 4th (highest) | 567 (16.1%) | 793 (28.6%) | | |
| Prescription Medications | | | 0.37 | <0.001 |
| 1st quartile (lowest) | 1,546 (43.9%) | 795 (28.6%) | | |
| 2nd | 1,090 (31.0%) | 921 (33.2%) | | |

| | | | | |
|---|---|---|---|---|
| 3rd | 642 (18.2%) | 671 (24.2%) | | |
| 4th (highest) | 240 (6.8%) | 389 (14.0%) | | |
| Baseline Costs, $ per member per year | | | | |
| Total | 7,154 (17,151) | 13,387 (25,827) | -0.28 | <0.001 |
| Inpatient | 466 (4,831) | 1,385 (9,374) | -0.12 | <0.001 |
| Outpatient | 5,049 (13,111) | 9,461 (20,102) | -0.26 | <0.001 |
| Prescription medications | 1,638 (7,891) | 2,541 (10,117) | -0.10 | <0.001 |
| Year of index date | | | 0.07 | |
| 2020 | 21 (0.6%) | 17 (0.6%) | | |
| 2021 | 56 (1.6%) | 54 (1.9%) | | |
| 2022 | 491 (14.0%) | 328 (11.8%) | | |
| 2023 | 575 (16.3%) | 482 (17.4%) | | |
| 2024 | 2,126 (60.4%) | 1,695 (61.1%) | | |
| 2025 | 249 (7.1%) | 200 (7.2%) | | |
| Starting BMI | | | 0.33 | |
| <18.5 | 0 (0.0%) | 0 (0.0%) | | |
| 18.5-24.9 | 86 (3.0%) | 7 (0.4%) | | |
| 25.0-29.9 | 480 (16.9%) | 209 (11.0%) | | |
| 30.0-34.9 | 1,453 (51.1%) | 918 (48.3%) | | |
| 35.0-39.9 | 824 (29.0%) | 765 (40.3%) | | |
| >40.0 | 0 (0.0%) | 0 (0.0%) | | |
| Ketone levels | | | 0.16 | <0.001 |
| 0-0.3 | 622 (25.5%) | 638 (31.7%) | | |
| 0.3-0.5 | 850 (34.9%) | 713 (35.4%) | | |
| 0.5+ | 967 (39.6%) | 664 (33.0%) | | |
| Time enrolled in INT | | | 0.06 | 0.19 |
| <6m | 1,126 (32.0%) | 919 (33.1%) | | |
| 6 to 12m | 1,239 (35.2%) | 943 (34.0%) | | |
| 12 to 24m | 802 (22.8%) | 670 (24.1%) | | |
| 24m plus | 351 (10.0%) | 244 (8.8%) | | |

Appendix 10. DID estimates for pooled sample

| Outcome | One year | | Two years | | Baseline means | | p-val, baseline trend difference |
|---|---|---|---|---|---|---|---|
| | DID Estimate | 95% CI | DID Estimate | 95% CI | UC | INT | |
| *Cost, $ per member per month* | | | | | | | |
| Total | -276.98*** | (-360.9, -193.06) | -242.73*** | (-322.29, -163.18) | 953.00 | 1078.08 | 0.90 |
| Inpatient | -110.82*** | (-154.56, -67.08) | -95.00*** | (-135.42, -54.58) | 114.31 | 112.39 | 0.90 |
| Outpatient | -46.57 | (-104.16, 11.02) | -43.11 | (-96.15, 9.93) | 461.43 | 563.01 | 0.58 |
| All Prescription Medications | -119.59*** | (-149.93, -89.25) | -104.62*** | (-136.82, -72.43) | 377.26 | 402.68 | 0.50 |
| *Utilization, visits per 1,000 members per month* | | | | | | | |
| Inpatient | -3.9*** | (-5.3, -2.6) | -3.5*** | (-4.8, -2.3) | 4.6 | 4.4 | 0.29 |
| ED | -2.1 | (-4.6, 0.4) | -1.9 | (-4.3, 0.5) | 24.6 | 21.9 | 0.07 |
| PCP | -66.6*** | (-76, -57.2) | -56.1*** | (-65.1, -47.1) | 236.9 | 266.5 | 0.04 |
| Cardiologist | -1.4 | (-2.8, 0.1) | -0.8 | (-2.2, 0.5) | 8.0 | 5.9 | 0.53 |
| Endocrinologist | -3.4*** | (-4.7, -2.1) | -2.6*** | (-3.8, -1.3) | 8.7 | 7.5 | 0.70 |

**Note:** *** $p < 0.001$, ** $p < 0.01$, * $p < 0.05$. The table reports coefficients, standard errors, and p-values for the interaction terms between the post-intervention indicator and the indicator for INT participation. Estimates are derived from a regression model of individual monthly outcomes adjusted for individual and calendar-month fixed-effects. Standard errors are clustered at the member level. Total cost is defined as the sum of inpatient, outpatient, and prescription medication costs. The baseline trend difference is the interaction term between INT participation and a linear month variable (1 through 12) estimated using baseline year data only. A $\chi^2$ test of the joint significance of the baseline trend difference was estimated for all cost outcomes and all utilization outcomes.

**Abbreviations:** DID, differences in differences; PMPM, per member per month; T2D, type 2 diabetes; ED, emergency department; PCP, primary care physician
